# A Randomized Double-Blind Placebo-Controlled Phase I/II Clinical Trial of a Human Papillomavirus Therapeutic Vaccine, PepCan, for Reducing Head and Neck Cancer Recurrence

**DOI:** 10.64898/2026.01.09.26343801

**Authors:** Emily Bivens, Omar Atiq, Teresa Evans, Milan Bimali, Ginger Brown, Jasmine Crane, Nadia Darwish, Jennifer L. Faulkner, Rangaswamy Govindarajan, Aaron Johnson, Alongkorn Kurilung, Oxana Lazarenko, Yong-Chen Lu, Keanna Marsh, Mauricio Moreno, Intawat Nookaew, Michael Robeson, Jumin Sunde, David Ussery, Emre Vural, Mindy Wilman, Mayumi Nakagawa

**Author notes:** Corresponding author: Mayumi Nakagawa, M.D., PhD., Department of Pathology, College of Medicine, University of Arkansas for Medical Sciences, 4301 West Markham Street Slot 502, Little Rock, AR 72205, U.S.A.

## Abstract

**Objectives:** Head and neck cancer (HNC) has a high recurrence rate. Safety and effectiveness of PepCan in reducing recurrence for HNC patients were assessed.

**Methods and analysis:** PepCan consists of four human papillomavirus 16 (HPV 16) E6 peptides and a *Candida* skin testing reagent (Candin®, Nielsen Biosciences) as a vaccine adjuvant. Since *Candida* was known to have a general immune stimulating effects, patients were recruited regardless of their HPV status. Men and women with HNC who had no evidence of disease after standard surgery, chemotherapy, and/or radiation treatments were enrolled. They were randomized at 3:1 to PepCan versus placebo. Seven intradermal injections of PepCan or placebo (saline) were given every 3 weeks (first 4 injections) or 3 months (last 3 injections). They were followed with two visits 6 months apart. Safety was assessed using Common Terminology Criteria for Adverse Events version 5, and efficacy was assessed based on not having recurrence within 2 years. In addition, immune responses were examined using enzyme-linked immunospot assay for HPV 16 E6 response, fluorescent-activated cell sorter analysis for peripheral immune cells, and T cell repertoire analysis. Peripheral cytokines and gut and oral microbiome were also analyzed.

**Results:** Seventeen patients were enrolled. The most common adverse events were grades 1 and 2 injection site reactions, and they occurred more frequently in the PepCan group (*p*<0.0001). Two patients had allergic reactions (grade 2 and grade 3), at the 6th vaccination, which were considered to be a dose-limiting toxicity (DLT). No serious adverse events were reported. In the intention-to-treat analysis (ITT), 45% (5/11) had non-recurrence in the PepCan group while 80% (4/5) had non-recurrence in the placebo group. For the per-protocol (PP) analysis, non-recurrence was 56% (5/9) for PepCan and 80% (4/5) for placebo. These differences were not statistically significant. Those who received PepCan and experienced non-recurrence had higher new T cell immune responses to HPV 16 E6 (*p*=0.05 for ITT and *p*=0.02 for PP). Pre-vaccination T helper type 1 cells were higher in the PepCan non-recurrence group compared to the PepCan recurrence group (*p*=0.01 for ITT and PP).

**Conclusions:** PepCan is safe although DLT can occur after multiple injections of PepCan. PepCan does not seem to be effective in reducing recurrence; however, the results are inconclusive given the small patient numbers.

What is already known on this topic

- Head and neck cancer (HNC) has a high recurrence rate after reaching no evidence of disease status after standard therapies including chemotherapy, radiation, immunotherapy and survey. However, no intervention is available to reduce recurrence.

What this study adds

- A therapeutic human papillomavirus vaccine called PepCan was tested in a clinical trial and has been shown to be safe.

How this study might affect research, practice or policy

- If a sufficiently powered study demonstrates efficacy in reducing recurrence rate, then how HNC patients are treated after achieving the no evidence of disease status will change.

## BACKGROUND

The estimated new cases of head and neck cancer (HNC) in the United States are 59,660, and the estimated deaths are 12,770 in 2025.^1^ Worldwide, the estimated new cases are 501,968 (188,960 for larynx, 120,416 for nasopharynx, 106,316 for oropharynx, and 86,276 for hypopharynx), and estimated deaths are 269,877 (103,216 for larynx, 73,476 for nasopharynx, 52,268 for oropharynx, and 40,917 for hypopharynx).^2^ Human papillomavirus (HPV) is best known as the causative agent of cervical cancer, but it can also cause cancers at other mucosal sites, including the anus, penis, vagina, vulva, and HNC. Incidences of HPV-associated HNC and anal cancers have increased notably.^3^ The recurrence rate of HNC is approximately 80% over 2 years (50%–60% local recurrence, 20%–30% distant recurrence).^4,5^

PepCan is a candidate therapeutic HPV vaccine our group developed, and it consists of HPV type 16 E6 peptides and a *Candida* adjuvant (Candin®, Nielson BioSciences, San Diego, CA). This *Candida* is a clear extract and not heat-killed. It is known to have general immune stimulation effects and has been shown to promote interleukin-12 (IL-12) secretion *in vitro* by Langerhans cells, which is mediated by dectin-1.^6^ *Candida* injection has been shown to be effective in regressing common warts.^7,8^ HPV transformation of squamous epithelium to a malignant phenotype is mediated by two early gene products, E6 and E7. ^9^ Both viral proteins interact with products of cellular human tumor-suppressor genes:^9^ the E6 protein can bind and promote degradation of cell-encoded p53, and the E7 protein interacts with the retinoblastoma susceptibility gene product.^10–12^ Expression of E6 and E7 proteins has been shown to be necessary and sufficient for HPV-16 transformation of human cells.^13–15^ While both proteins are possible candidates as components of therapeutic HPV vaccine, we chose to include E6 because T cell immune responses to E6 but not E7 has been associated with regression of cervical intraepithelial neoplasia (CIN).^16,17^ PepCan was first tested for its ability to regress CIN.^18,19^ In this study, we examined its potential to reduce recurrence of HNC.

## METHODS

### PATIENTS

Recruitment for this study took place between October 28, 2019 and June 29, 2023. Inclusion criteria were being men or women aged 18 years or older, having completed curative therapy (surgery and/or radiation and/or chemotherapy) within the previous 120 days for squamous cell carcinoma of the head and neck, and having achieved no evidence of disease based on clinical and/or radiographic evaluations. Since *Candida* was known to have a general immune stimulating effects, patients were enrolled regardless of their HPV status.^6^ After signing a written informed consent, patients (n=21) were screened, and those who met the inclusion criteria and returned a stool sample (n=17) were eligible for vaccination. They were randomly assigned to the PepCan or placebo (saline) arm at a 3:1 ratio as shown in a Consort diagram (**Figure 1**). Exclusion criteria included a positive urine pregnancy test, being pregnant or attempting to become pregnant, breastfeeding, allergy to *Candida*, a history of severe asthma, having previously received PepCan, or a history of recurrent HNC. A full description of eligibility criteria is in the trial protocol submitted with this manuscript. The patients and the public did not provide input for this study. The study adhered to the Declaration of Helsinki and was prospectively registered at ClinicalTrials.gov (NCT03821272).

**Figure 1.**
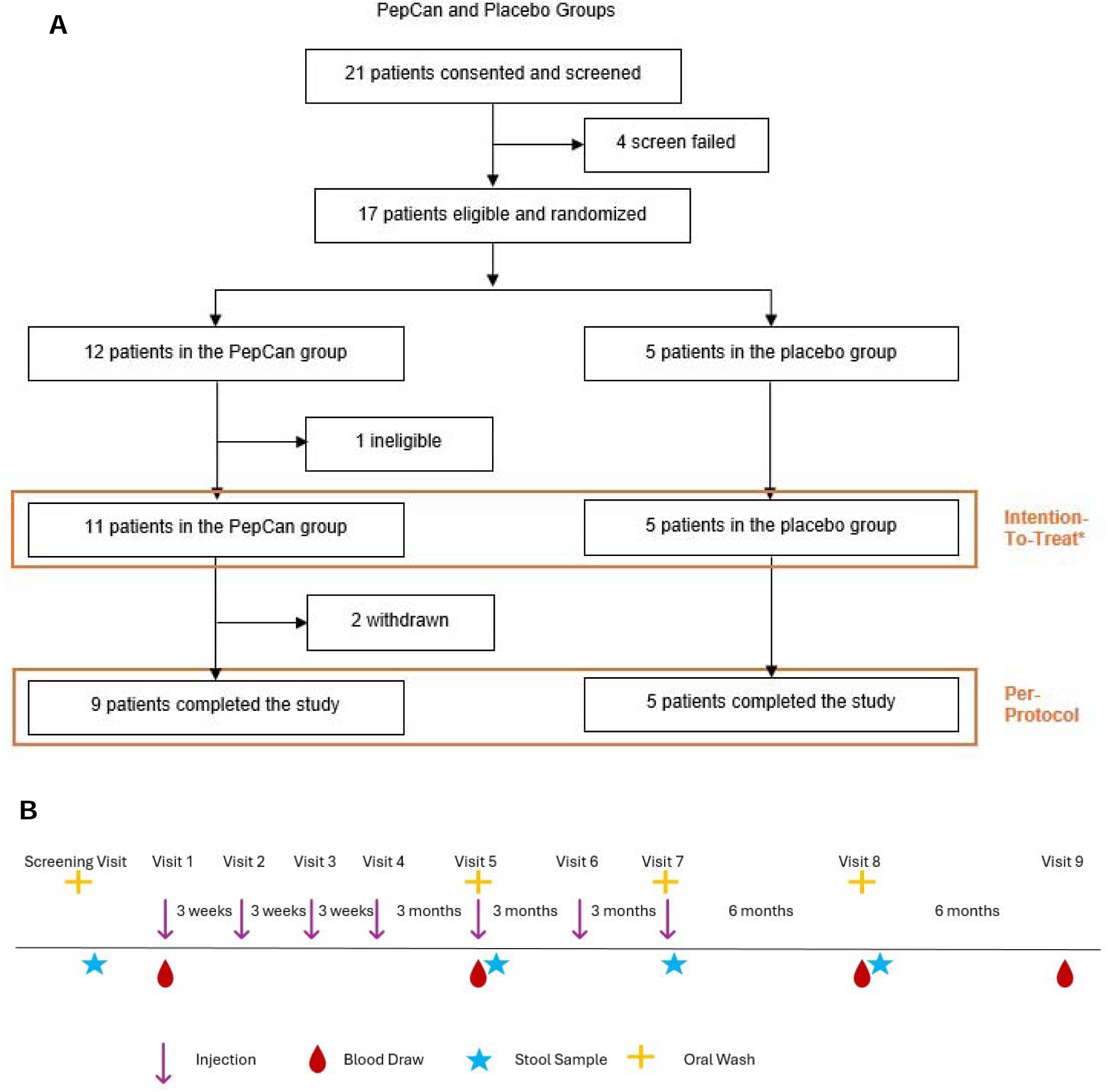
Consort diagram and the trial design. **A.** A consort diagram summarizing patient progress during the trial. One patient in the PepCan arm was found to be “ineligible” for having had a previous recurrence. This patient was followed for safety but excluded from efficacy analysis. **B.** The first 4 injections (PepCan or placebo) were given every three weeks, and the next 3 injections were given every 3 months. Patients had two follow-up visits six months apart after the final injection. Blood was collected at visits 1, 5, 8, and 9 for immunological assessments by ELISPOT, bulk TCR deep sequencing, and FACS analysis, and for cytokine analysis. Oral wash samples were collected at screening, visit 5, visit 7, and visit 8 and stool samples were collected at home after these visits for microbiome analyses.

### VACCINE COMPOSITION AND DELIVERY

PepCan consists of four current good manufacturing production-grade synthetic peptides covering the HPV 16 E6 protein [amino acid (aa)1-45, 46-80, 81-115, and 116-158] (CPC Scientific, San Jose, CA; Curia, Albany, NY). Lyophilized peptides (50 μg per peptide) were reconstituted with sterile water and mixed in a syringe with 0.3 mL of *Candida* (Candin®, Nielsen Biosciences, San Diego, CA). Candin is an extract of *Candida*, and is not heat-killed nor fixed. The PepCan mixture was administered intradermally in any limb, but most often in the anterior forearm. For the placebo arm, an equal volume of normal saline was administered.

### TRIAL DESIGN AND END POINTS

This clinical trial (University of Arkansas for Medical Sciences Institutional Review Board approval #217672) was a randomized, double-blind, placebo-controlled, single-center, Phase I/II study in which patients were assigned to PepCan or placebo at a 3:1 ratio using a computer-generated randomization scheme created by the study statistician. The patients and all study personnel were blinded to which vaccine the patients received, except for the research pharmacists who assigned patients to interventions. Only the study statistician and the study pharmacists had access to the randomization scheme. At the screening visit, the patients were given a questionnaire which asked whether they received HPV prophylactic vaccination, their motivations for participation, employment, education, number of children, number of lifetime sexual partners, type of sexual activities, and use of alcohol, tobacco or other nicotine products. After enrollment, the first four injections were given 3 weeks apart while the last three injections were given 3 months apart. Then, the patients returned for observational visits 6 and 12 months after the last injection. The primary endpoint was safety, which was assessed from the time of informed consent until visit 9 or until recurrence according to the National Cancer Institute Common Terminology Criteria for Adverse Events (CTCAE) Version 5. The dose-limiting toxicities (DLTs) were defined as grade 2 or above allergic/autoimmune reactions and any grade 3 or above adverse events (AEs).The secondary endpoint was clinical response, which was assessed by lack of recurrence within two years.

## IMMUNOLOGICAL ASSESSMENTS

### ELISPOT Assay

Peripheral immune responses were examined by interferon-L enzyme-linked immunospot (ELISPOT) assay, bulk T cell receptor (TCR) deep sequencing, and immune profiling. T cell lines were established from blood draws from visits 1, 5, 8, and 9 as described previously.^20^ In short, peripheral blood mononuclear cells (PBMCs) were isolated from heparinized whole blood using a Ficoll density-gradient centrifugation method, separated into CD14+ monocytes and CD14-depleted PBMCs, and cryopreserved. Autologous dendritic cells were established by growing monocytes in the presence of granulocyte monocyte-colony stimulating factor (50 ng/mL, Sanofi-Aventis, Paris, France) and recombinant interleukin-4 (100 U/mL, R&D Systems, Minneapolis, MN) for seven days, and were matured by 48-hour culture in wells containing irradiated mouse L-cells expressing CD40 ligands. CD3 T cells were magnetically selected (Pan T Cell Isolation Kit II, Miltenyi Biotec, Auburn, CA) from CD14-depleted PBMCs. Human papillomavirus type 16 (HPV 16) E6- and E7-specific CD3 T cell lines were established by *in vitro* stimulation of CD3 cells for seven days with autologous dendritic cells pulsed with E6-glutathione *S*-transferase and E6 expressing recombinant vaccinia virus or E7-glutathione S-transferase and E7 expressing recombinant vaccinia virus.^21,22^ *In vitro* stimulation was repeated for an additional seven days.

ELISPOT assays were performed in triplicate using overlapping peptides covering the ten regions (aa1-25, 16-40, 31-55, 46-70, 61-85, 76-100, 91-115, 106-130, 121-145, and 136-158) within the E6 protein of HPV 16, as previously described.^20^ Briefly, 96-well plates (MultiScreen-MSIPS 4510 plates, Sigma-Aldrich, St. Louis MO) were treated with 35% ethanol in sterile phosphate-buffered saline and rinsed immediately. They were subsequently coated with 5 μg/ml of primary anti-interferon-L monoclonal antibody (1-D1K, Mabtech, Sweden), washed, and blocked. Then, 2.5 x 10^4^ CD3 T cells were added per well in triplicate along with pooled peptides (10 μM per peptide). Each peptide pool contained 3 overlapping HPV 16 E6 15-mer peptides. For example, the E6 1-25 pool contained E6 1-15, E6 6-20, and E6 11-25 peptides. Negative control wells contained media only, and positive control wells contained phytohaemagglutinin at 10 μg/mL, (Remel, Lenexa, KS). Plates were incubated for 24 hours at 37°C with 5% carbon dioxide. After washing, a secondary biotin-conjugated anti-Interferon-L monoclonal antibody (7-B6-1, Mabtech, Nacka Strand, Sweden) was added at 1 µg/ml and incubated for 2 hours. After washing, streptavidin-horseradish peroxidase (Mabtech) was added at a dilution of one to 750 and incubated at room temperature for one hour. Wells were washed and coated with 3,3’,5,5’ tetramethylbenzidine (Mabtech) for 10 to 30 minutes and allowed to develop at room temperature until spots emerged. The plates were then rinsed in water and allowed to dry overnight. Peptide pools with spot forming units twice or greater to the media only control were considered to be positive. The ratio between spot forming units in peptide wells to the spot forming units in the media control is called positivity index.

### Fluorescent-activated cell sorter (FACS) analysis

Immune profiling measuring T-helper type 1(Th1) cells, T-helper type 2 (Th2) cells, and regulatory T cells (Tregs), was also performed using FACS with blood samples drawn at visits 1, 5, 8, and 9 using previously established methods^18,19^ with minor modifications. To analyze Th1 cells (CD4- and Tbet-positive), Th2 cells (CD4- and GATA3-positive), and Tregs (CD4-, CD25-, and FoxP3-positive), thawed PBMCs were treated with Fc Receptor Blocker (Innovex, Maple Plain, MN) and stained using fluorescein isothiocyanate-labeled anti-human CD4 (Thermo Fisher Scientific, Hillsboro, OR), phycoerythrin-labeled anti-human/mouse T-bet (Thermo Fisher Scientific), allophycocyanin -eFluor-labeled anti-human CD25 (clone BC96, Thermo Fisher Scientific), allophycocyanin-labeled anti-human Foxp3 (Thermo Fisher Scientific), and phycoerythrin -Cy7 labeled anti-human/mouse GATA3 (BD, Franklin, NJ). Cells were initially stained with antibodies for surface markers CD3, CD4, and CD25. Intracellular staining for T-bet, GATA3, and Foxp3 was performed using the Foxp3 staining kit (Thermo Fisher Scientific). Live/dead staining 7-AAD (BD) was used to exclude dead cells. Stained cells were analyzed using FACSCelesta (BD) available in the University of Arkansas for Medical Sciences Microbiology and Immunology Flow Cytometry Core Laboratory, and FlowJo (BD) was used to analyzed the data.

### TCR repertoire analysis

Bulk deep sequencing of polymerase chain reaction (PCR)-amplified TCR β CDR3 regions using genomic DNA from PBMCs was performed (Adaptive Biotechnologies, Seattle, WA).^23^ Using bias-controlled V and J gene primers, the rearranged V(D)J segments were amplified and sequenced. A clustering algorithm was used to correct for sequencing errors, and the V(D)J segments were annotated according to the International ImMunoGeneTicsCollaboration^24,25^ to identify the V, D, and J genes that contributed to each rearrangement. A mixture of synthetic TCR analogs was used in PCR to estimate the number of cells bearing each unique TCR sequence.^26^ Putatively vaccine-specific T cells were identified by comparing post-vaccination PBMC sample to the pre-vaccination PBMC sample using a beta-binomial model^27^ under the immunoSeq analyzer (Adaptive Biotechnologies). The results were obtained for the numbers of clonotypes as well as the fractions of total T cells.

### CYTOKINE ASSESSMENTS

To identify potential biomarkers predictive of vaccine response and to examine the effects of vaccination, plasma samples for 81 plasma cytokines were analyzed using methods described previously.^18^ As our pilot data demonstrated that approximately one third of cytokine levels were significantly different when the blood samples were processed at 2 hours versus 1 hour,^18^ we used plasma samples that were processed within 1 hour of blood draw. The quantities of β-nerve growth factor (β-NGF), cutaneous T cell-attracting chemokine (CTACK), eotaxin, basic fibroblast growth factor (FGF basic), granulocyte-colony stimulating factor (G-CSF), granulocyte monocyte-colony stimulating factor (GM-CSF), C-X-C motif chemokine ligand 1 (CXCL1, also known as GRO-α), hepatocyte growth factor (HGF), interferon-α2 (IFN-α2), interferon-γ (IFN-γ), interleukin-1α (IL-1α), interleukin-1β (IL-1β), IL-1 receptor agonist (IL-1RA), interleukin-2 (IL-2), interleukin-2Rα (IL-2Rα), interleukin-3 (IL-3), interleukin-4 (IL-4), interleukin-5 (IL-5), interleukin-6 (IL-6), interleukin-7 (IL-7), interleukin-8 (IL-8), interleukin-9 (IL-9), interleukin-10 (IL-10), interleukin-12 (IL-12) (p70), IL-12 (p40), interleukin-13 (IL-13), interleukin-15 (IL-15), interleukin-16 (IL-16), interleukin-17A (IL-17A), interleukin-18 (IL-18), IFN-γ-induced protein 10 (IP-10), leukemia inhibitory factor (LIF), monocyte-colony stimulating factor (M-CSF), monocyte chemotactic protein 1 (MCP-1 or MCAF), monocyte chemotactic protein 3 (MCP-3), macrophage migration inhibitory factor (MIF), C-X-C motif chemokine ligand 9 (CXCL9, also known as MIG), macrophage inflammatory protein1α (MIP-1α), macrophage inflammatory protein-1β (MIP-1β), platelet-derived growth factor subunit B (PDGF-BB), regulated on activation, normal T-cell expressed and secreted (RANTES), stem cell factor (SCF), stem cell growth factor β (SCGF-β), stromal-derived cell factor-1α (SDF-1α), tumor necrosis factor α (TNF-α), tumor necrosis factor β (TNF-β), tumor necrosis factor-related apoptosis inducing ligand (TRAIL), and vascular endothelial growth factor (VEGF) were determined using a commercially available Bio-Plex 48-plex kit (Bio-Rad Laboratories) according to the manufacturer’s instructions using a Bio-Plex 200 instrument (Bio-Rad Laboratories). The quantities of a proliferation-inducing ligand (APRIL/TNSFS13), B-cell activating factor (BAFF/TNFSF13B), soluble CD30 (sCD30/TNFRSF8), soluble CD163 (sCD163), chitinase-3-like 1, glycoprotein 130/soluble interleukin-6 receptorβ (gp130/sIL-6Rβ), interferon-β (IFN-β), soluble interleukin-6 receptorα (sIL-6Rα), interleukin-11 (IL-11), interleukin-19 (IL-19), interleukin-20 (IL-20), interleukin-22 (IL-22), interleukin-26 (IL-26), interleukin-27 (IL-27, p28), interleukin-28A/interferon-λ2 (IL-28A/IFN-λ2), interleukin-29/interferon-λ1 (IL-29/IFN-λ1), interleukin-32 (IL-32), interleukin-34 (IL-34), interleukin-35 (IL-35), tumor necrosis factor superfamily member 14 (LIGHT), matrix metalloproteinase-1 (MMP-1), MMP-2, MMP-3, osteocalcin, osteopontin (OPN), pentraxin-3, soluble tumor necrosis factor-receptor 1 (sTNF-R1), sTNF-R2, thymic stromal lymphopoietin (TSLP), and tumor necrosis factor-like weak inducer of apoptosis (TWEAK/TNSFS12) were determined separately using another Bio-Plex inflammation kit. Transforming growth factor-β1 (TGF-β1), transforming growth factor-β2 (TGF-β2), and transforming growth factor-β3 (TGF-β3) were determined separately using Bio-Plex TGF-β kit which included an acid treatment.

### MICROBIOME ASSESSMENTS

Oral wash samples were collected in clinic at screening visit and visits 5, 7, and 8, and fecal samples were collected by patients at home using the OMNIgene.GUT tubes (DNA Genotek, USA) following the same visits. The oral wash samples were collected using 10 mL of mouth wash which was swished in mouth for 30 seconds. DNA was extracted using ZymoBIOMICS DNA Miniprep Kit (Zymo Research, Irvine, CA) and was aliquoted. Stool DNA samples were analyzed using a flow cell (R10.4.1) on a MinION Mk1B sequencer (Oxford Nanopore Technologies, New York, NY) and amplicon-based rRNA sequencing (Argonne National Laboratory, Lemont, IL). Oral wash DNA was analyzed using the amplicon-based method only due to high human DNA content.

A positive control utilized in this study was mock vaginal microbial communities composed of a mixture of genomic DNA from the American Type Culture Collection (Manassas, VA) and Zymo Research. The negative control was OMNIgene.GUT preservation solution without the sample as blank extraction.^28^ Controls and the extracted DNA were amplified and analyzed using the 16S rRNA gene on an Illumina MiSeq sequencing platform.^29^ The same volume of DNA was used for each reaction, and then normalized at the PCR pooling step. This ensures that equal amounts of each amplified sample are added to the sequencing pool. Paired-end reads from libraries with ∼250-bp inserts were generated for the V4 region using the barcoded primer set: 515FB: 5’-GTGYCAGCMGCCGCGGTAA-3’ and 806RB: 5’-GGACTACNVGGGTWTCTAAT-3’.^30–34^ MiSeq Reagent Kit v2 (2 × 150 cycles, MS-102-2002) was used. Initial sequence processing and analyses were performed using QIIME 2.^35^

### TRIAL OVERSIGHT

This clinical trial was supported by the National Institutes of Health. No commercial support was available for this study. All authors participated in data acquisition, had access to data, wrote and/or edited the manuscript, and confirmed the accuracy and completeness of the data.

### STATISTICAL ANALYSIS

The recurrence rate of head and neck cancer is 80% over 2 years (50 to 60% local recurrence and 20 to 30% distant recurrence).^4,5^ In the Phase I trial of PepCan described above the overall histological regression rate of HSILs was 45%.^18^ A reduced recurrence rate by the magnitude of 45% would reach 44%. Vaccinating 72 subjects in the PepCan arm and 24 subjects in the placebo arm would have 90% power at two-sided alpha of 5% based on Fisher’s exact test. Accounting for about 5% drop out rate, 75 subjects would be needed in the PepCan arm, and 25 subjects in the placebo arm. The difference between PepCan non-recurrence and PepCan recurrence and between the PepCan and the placebo groups were assessed using a Fisher’s exact test. The 95% confidence intervals were computed using a Clopper-Pearson method. The difference between the injection-related AEs for PepCan and placebo groups were assessed also using the Fisher’s exact test. A two-sided *p*-value of less than 0.05 was used to determine statistical significance, and corrections for multiple comparisons were made where appropriate. A more detailed descriptions of statistical methods are described in the Statistical Analysis Plan.

## RESULTS

### PATIENTS AND TREATMENT

Twelve patients were randomly assigned to the PepCan arm and five patients were assigned to the placebo (saline) arm (**Figure 1**). One patient assigned to the PepCan arm was determined to be ineligible during the study due to a previous recurrence, and this individual was included in the baseline characteristics and followed for the remainder of the study for safety. This patient was excluded from efficacy analysis. Remaining patients in the PepCan and placebo arms were included in the ITT population (n=16) including two withdrawn patients who were considered to be non-responders (i.e., recurrence). These withdrawn patients were excluded form PP population (n=14).

The screening questionnaire (**Figure S1**) revealed that none (out of 18 patients who completed the questionnaire including one screen failure) had ever received an HPV prophylactic vaccine. The main motivation for participating was to take care of personal health for 66.7% (12/18). Most patients are either full-time workers (27.8% or 5/18), retired (27.8% or 5/18), or disabled (27.8% or 5/18). The majority’s highest education received was either high school (33.3% or 6 /18) or enrolled but did not complete college (33.3% or 6/18). Most commonly, they had 3 to 5 children (44.4% or 8/18). The number of lifetime sexual partners ranged from 1 to 50 with 44.4% (8/18) having 5 to 9 lifetime partners being most common. Twelve patients (66.7%) have reported participating in vaginal sex, four (22.2%) did not want to say, and one (5.6%) reported not having sex. Ten (55.6%) were former but not current tobacco users, and five (27.8%) were current users. Eleven (61.1%) did not drink alcohol on a regular basis while seven did (38.9%).

Their baseline characteristics were mostly well balanced, but the placebo group included only patients with squamous cell carcinoma sites of the pharynx while the PepCan group included sites of the pharynx, larynx, and oral (**Table 1**). The patient in the placebo group mostly had T2 disease, but the patients in the PepCan group were evenly distributed among T1 through T4 diseases. Enrollment was halted early after 17 patients were enrolled because of vaccine peptide production failure and because of the results of another clinical trial which showed more efficacy with *Candida* alone in comparison to PepCan.^20^

**TABLE 1.** Baseline characteristics of the patients.

| Characteristic | Overall<br>N = 17 | PepCan<br>N = 12 | Placebo<br>N = 5 |
| --- | --- | --- | --- |
| **Age** |  |  |  |
| Mean (SD) | 59.82 (9.82) | 60.03 (8.30) | 59.34 (14.00) |
| Median (IQR) | 61.40 (13.00) | 62.60 (9.55) | 54.30 (23.00) |
| (Min., Max.) | (41.30,74.80) | (41.30,69.20) | (43.80,74.80) |
| **Sex** |  |  |  |
| Male | 15/17 (88.2%) | 11/12 (91.7%) | 4/5 (80%) |
| Female | 2/17 (11.8%) | 1/12 (8.3%) | 1/5 (20%) |
| **Race** |  |  |  |
| White | 12 / 17 (71%) | 7 / 12 (58%) | 5 / 5 (100%) |
| Black | 4 / 17 (24%) | 4 / 12 (33%) | 0 / 5 (0%) |
| Other | 1 / 17 (5.9%) | 1 / 12 (8.3%) | 0 / 5 (0%) |
| **Ethnicity** |  |  |  |
| Non-Hispanic | 16 / 17 (94%) | 11 / 12 (92%) | 5 / 5 (100%) |
| Hispanic/Latino | 1 / 17 (5.9%) | 1 / 12 (8.3%) | 0 / 5 (0%) |
| **WBC** |  |  |  |
| Mean (SD) | 5.03 (1.55) | 5.20 (1.52) | 4.64 (1.70) |
| Median (IQR) | 5.04 (2.05) | 5.32 (1.69) | 4.33 (1.98) |
| (Min., Max.) | (2.96,8.24) | (2.96,8.24) | (3.00,7.22) |
| **Hemoglobin** |  |  |  |
| Mean (SD) | 11.63 (1.81) | 11.94 (1.91) | 10.88 (1.43) |
| Median (IQR) | 12.10 (2.50) | 12.30 (1.73) | 11.10 (2.70) |
| (Min., Max.) | (8.30,15.40) | (8.30,15.40) | (9.30,12.30) |
| **Hematocrit** |  |  |  |
| Mean (SD) | 35.55 (5.56) | 36.63 (5.73) | 32.96 (4.64) |
| Median (IQR) | 36.50 (8.60) | 37.50 (6.15) | 35.00 (8.90) |
| (Min., Max.) | (26.40,46.90) | (26.40,46.90) | (27.90,37.00) |
| **BMI** |  |  |  |
| Mean (SD) | 22.17 (3.87) | 22.30 (3.82) | 21.86 (4.41) |
| Median (IQR) | 22.65 (4.91) | 22.99 (4.51) | 20.67 (4.91) |
| (Min., Max.) | (15.82,29.25) | (15.82,29.25) | (16.46,27.76) |
| **HPV** |  |  |  |
| Negative | 1 / 1 (100%) | 0 / 0 (-%) | 1 / 1 (100%) |
| Unknown | 16 | 12 | 4 |
| **P16** |  |  |  |
| Positive | 7 / 12 (58%) | 4 / 9 (44%) | 3 / 3 (100%) |
| Negative | 5 / 12 (42%) | 5 / 9 (56%) | 0 / 3 (0%) |
| Unknown | 5 | 3 | 2 |
| **HPV/P16** |  |  |  |
| Positive | 7 / 13 (54%) | 4 / 9 (44%) | 3 / 4 (75%) |
| Negative | 6 / 13 (46%) | 5 / 9 (56%) | 1 / 4 (25%) |
| Unknown | 4 | 3 | 1 |
| **Site** |  |  |  |
| Pharynx | 11 / 17 (65%) | 6 / 12 (50%) | 5 / 5 (100%) |
| Larynx | 3 / 17 (18%) | 3 / 12 (25%) | 0 / 5 (0%) |
| Oral | 3 / 17 (18%) | 3 / 12 (25%) | 0 / 5 (0%) |
| **Tumor** |  |  |  |
| T1 | 3 / 17 (18%) | 3 / 12 (25%) | 0 / 5 (0%) |
| T2 | 6 / 17 (35%) | 2 / 12 (17%) | 4 / 5 (80%) |
| T3 | 4 / 17 (24%) | 3 / 12 (25%) | 1 / 5 (20%) |
| T4 | 4 / 17 (24%) | 4 / 12 (33%) | 0 / 5 (0%) |
| **Lymph node** |  |  |  |
| N0 | 2 / 17 (12%) | 1 / 12 (8.3%) | 1 / 5 (20%) |
| N2 | 10 / 17 (59%) | 8 / 12 (67%) | 2 / 5 (40%) |
| N1 | 3 / 17 (18%) | 1 / 12 (8.3%) | 2 / 5 (40%) |
| N3 | 2 / 17 (12%) | 2 / 12 (17%) | 0 / 5 (0%) |
| **Metastasis** |  |  |  |
| M0 | 17 / 17 (100%) | 12 / 12 (100%) | 5 / 5 (100%) |

### SAFETY

Injection-related commonly occurring AEs are summarized in **Table 2**, and all AEs, regardless of relatedness to treatments are summarized in **Table S1**. DLTs occurred in two patients at their 6^th^ visit vaccination with grade 2 and grade 3 allergic reactions respectively. They responded well to treatment with antihistamine and steroids. These patients did not receive the 7^th^ and final vaccination. The most common AE was grades 1 and 2 injection site reactions which occurred only with the PepCan group (23/72 injections) but not with the placebo group (*p*<0.0001). Most other AEs such as headache and vaccination complication occurred more commonly in the PepCan group, but not significantly.

**TABLE 2.** Injection-related adverse events.

|  |  | CTCAE v5 AE Severity Scale Grade |  |  |  |  |  |  |  |  |  |  |  |
| --- | --- | --- | --- | --- | --- | --- | --- | --- | --- | --- | --- | --- | --- |
|  |  | PepCan n=72 |  |  |  |  |  | Placebo n=35 |  |  |  |  |  |
| CTCAE v5 System Organ Class | CTCAE v5 AE Term | 1 | 2 | 3 | 4 | 5 | Total | 1 | 2 | 3 | 4 | 5 | Total |
| General Disorders and Site Administration Conditions | Injection site reaction | 20 | 3 | - | - | - | 23 | - | - | - | - | - | - |
|  | Fatigue | - | - | - | - | - | - | 1 | - | - | - | - | 1 |
| Immune System Disorders | Allergic reaction | - | 1 | 1 | - | - | 2 | - | - | - | - | - | - |
| Injury, Poisoning and Procedural Complications | Vaccination complication | 3 | 1 | - | - | - | 4 | 1 | - | - | - | - | 1 |
| Metabolism and Nutrition Disorders | Hypokalemia | 1 | - | - | - | - | 1 | - | - | - | - | - | - |
| Nervous System Disorders | Dizziness | 1 | - | - | - | - | 1 | - | - | - | - | - | - |
|  | Headache | 2 | - | - | - | - | 2 | - | - | - | - | - | - |
| Skin and Subcutaneous Tissue Disorders | Pruritus | - | 1 | - | - | - | 1 | - | - | - | - | - | - |

≥ grade 2 allergic reactions were considered to be dose-limiting toxicities. N, number of injections

**TABLE 3.** Outcomes.

| **Characteristic** | **Overall**<br>**N = 17** | **PepCan**<br>**N = 12** | **Placebo**<br>**N = 5** |
| --- | --- | --- | --- |
| **Outcome** |  |  |  |
| Non-recurrence | 9 / 17 (53%) | 5 / 12 (42%) | 4 / 5 (80%) |
| Recurrence | 5 / 17 (29%) | 4 / 12 (33%) | 1 / 5 (20%) |
| Withdrawn | 2 / 17 (12%) | 2 / 12 (17%) | 0 / 5 (0%) |
| Ineligible | 1 / 17 (5.9%) | 1 / 12 (8.3%) | 0 / 5 (0%) |
| **Outcome (ITT)<sup>1</sup>** |  |  |  |
| Non-Recurrence | 9 / 16 (56%) | 5 / 11 (45%) | 4 / 5 (80%) |
| Recurrence | 7 / 16 (44%) | 6 / 11 (55%) | 1 / 5 (20%) |
| **Outcome (PP)<sup>2</sup>** |  |  |  |
| Non-Recurrence | 9 / 14 (64%) | 5 / 9 (56%) | 4 / 5 (80%) |
| Recurrence | 5 / 14 (36%) | 4 / 9 (44%) | 1 / 5 (20%) |

Efficacy was assessed as “non-recurrence” within 2 years from beginning the study.
<sup>1</sup>The ITT analysis excluded an “ineligible” subject.
<sup>2</sup>The PP analysis included patients who completed the study, and excluded those who were “ineligible” or “withdrawn”.

### EFFICACY

Patients with non-recurrence over the ten visits (the screening and the 9 study visits over two years) were considered to be responders while patients with recurrence were considered as non-responders. Analysis of the ITT group revealed non-recurrence in 45% (5 /11) of patients in the PepCan group [95% confidence interval (CI), 0.17 to 0.77] and 80% (4/5) non-recurrence for the placebo group (95% CI, 0.28 to 0.99). Analysis of the PP group revealed non-recurrence in 56% (5/9) of the PepCan group (95% CI, 0.21 to 0.86) and non-recurrence in 80% (4/5) of the placebo group (95% CI, 0.28 to 0.99) Neither comparison was significant with *p*-values of 0.31 and 0.58, respectively. Unexpectedly, the placebo group showed higher non-recurrence rate; but the results are inconclusive given the small number of patients enrolled.

## IMMUNE RESPONSES

### ELISPOT assay

Representative results are shown in **Figure S2**. ELISPOT analysis revealed no difference between the percentage of patients who had anti-HPV 16 E6 response prior to vaccine in the PepCan and placebo arms. Likewise, no difference was found in response prior to vaccine for PepCan non-recurrence and recurrence groups. No difference was found in the development of a new response to HPV 16 E6 in at least one region between PepCan and placebo groups. However, significantly more patients in the PepCan non-recurrence group (5 of 5 for ITT and PP) developed a new response for HPV 16 E6 after vaccination compared to patients in the PepCan recurrence group [1/5 for ITT(*p*=0.05) and 0/3 for PP (*p*=0.02)].

### FACS analysis

Comparisons were made of pre-vaccination levels between PepCan and placebo groups and between PepCan non-recurrence and PepCan recurrence groups. The ITT analysis revealed significantly higher Th1 cells in the PepCan non-recurrence group compared to PepCan recurrence group pre-vaccination (**Figure 2B**, *p*=0.01). The results were similar for the PP analysis (**Figure S3B**, *p*=0.04). When these groups were compared across visits 1 (pre-vaccination), 5, 7, and 8, some significant changes were noted but not consistently (**Figure 2** panels C and D for ITT and **Figure S2** panels C and D for PP).

**Figure 2.**
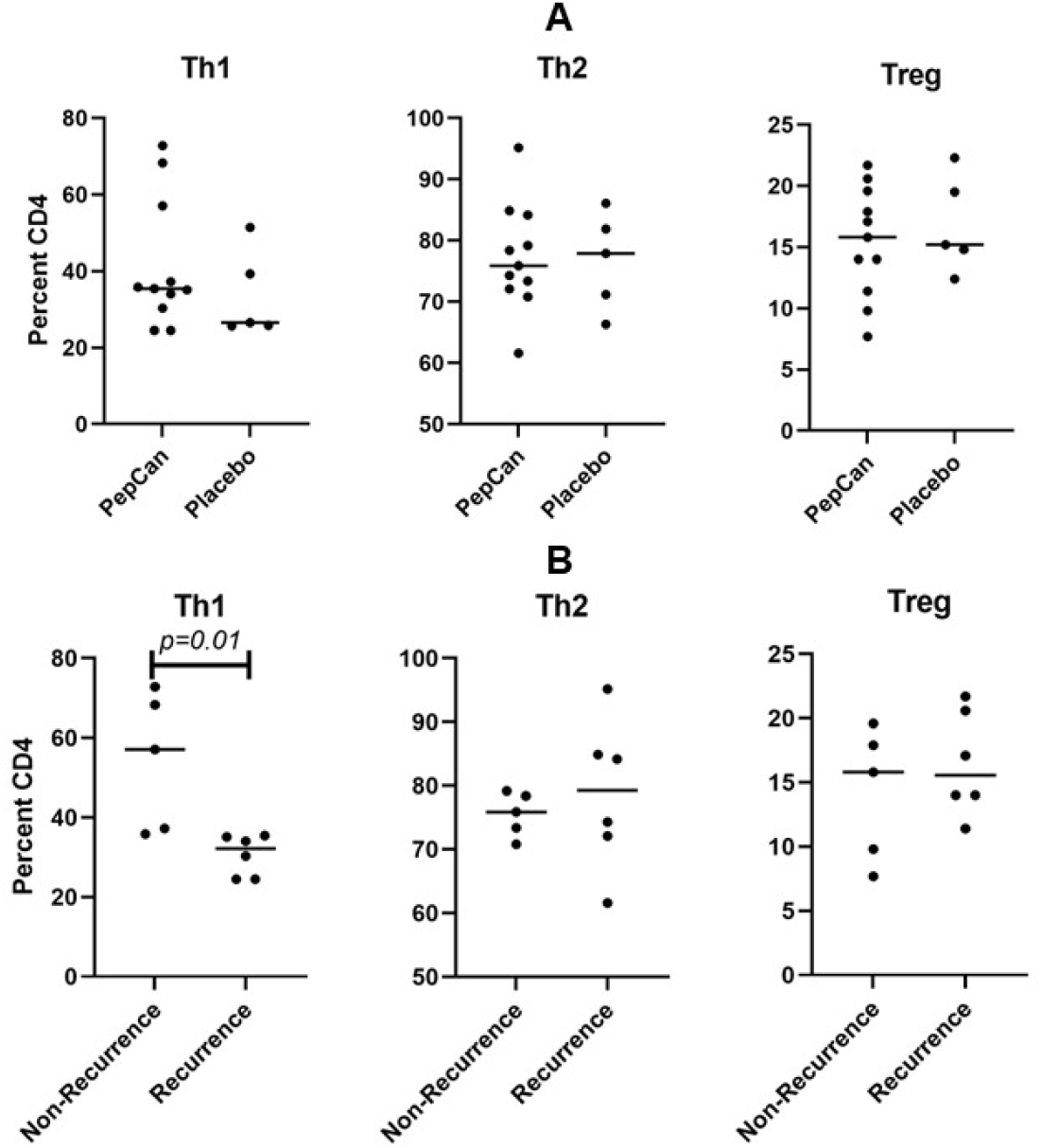
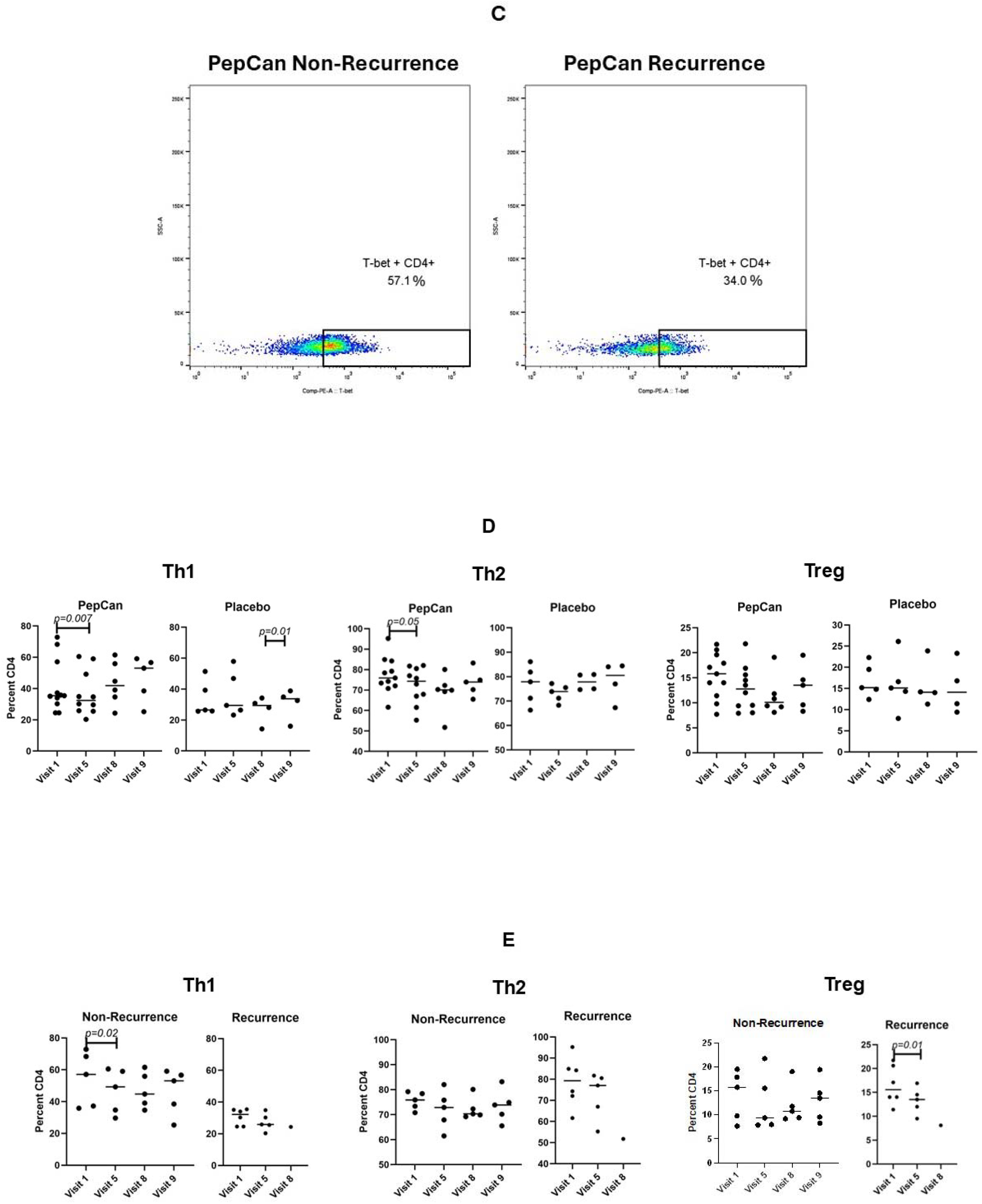
FACS analyses of peripheral immune cells ITT. **A.** Analyses of Th1, Th2, and Treg at visit 1 (pre-vaccination) between the PepCan and placebo groups. **B**. Analyses of Th1, and Th2, and Treg also at visits 1 between the PepCan non-recurrence and the PepCan recurrence groups. **C**. Representative raw data plots for Th1 results for a PepCan non-recurrence (Patient 17) and a PepCan recurrence (Patient 11) patients. **D**. Analyses of Th1, and Th2, and Treg also at visits 1, 5, 8, and 9 between the PepCan and placebo groups. **E**. Analyses of Th1, Th2, and Treg between non-recurrence PepCan group and recurrence PepCan group at visits 1, 5, 8, and 9.

### TCR repertoire analysis

Comparisons between PepCan and placebo groups and between PepCan non-recurrence and PepCan recurrence groups were made in ITT (**Figure S4** panels A-D) and PP (panels E-H) populations. Data are presented as the total number of templates (i.e., the total number of putatively vaccine-specific T cells; **Figures S4** panels A, B, E, and F) and as a fraction of total cells (**Figures S4** panels C, D, G, and H). No differences were found.

### CYTOKINE ANALYSIS

In the ITT analysis, the mean IL5 level was significantly lower in the PepCan group compared to placebo group (**Figure S5A**) (Difference, -227.91; 95% CI, -372.88 to -82.95; *p* < 0.01). For MCP1_MCAF and SDF1a, the interaction effect was retained suggesting that the treatment effect was moderated by study visit. The mean MCP1-MCAF level was significantly lower in the PepCan group compared to the placebo group for visit 1 (Difference, -34.38; 95% CI: -56.76 to -12.00; *p* < 0.01). Other significant changes in SDF1a and BAFF_TNFSF13B among placebo-treated patients were found for some visit comparisons (**Figure S5A**).

As to the comparisons between the PepCan non-recurrence and recurrence group ITT, the mean IL5 level was significantly higher among the PepCan non-recurrent patients (**Figure S5B**) (Difference, 215.49; 95% CI, -86.81 to 344.17; *p* < 0.01). The interaction effect was retained for IL1ra, IL12_p40, and BAFF_TNFSF13B for some visit comparisons (**Figure S5B**).

For PP analysis comparing PepCan and the placebo groups, the only significance retained was that for MCP1_MCAF. When comparing PepCan non-recurrence and recurrence groups, the mean chitinase-3-like 1 level was significantly lower at visit 5 compared to visit 1 (Difference, -3457.98; 95% CI, -5401.79 to -1514.16; *p* < 0.01), and the interaction effect was retained for IL28A_IFNI2 and MMP1 (Data not shown).

## MICROBIOME

### Stool

The compositions of stool bacteria at phylum and species are level are shown in **Figure S6A** and **Figure S6B** respectively for metagenomics. Since more diverse fecal microbiome has been associated with better outcomes for cancer patients treated with checkpoint inhibitors,^36–41^ we compared alpha diversity between PepCan and placebo groups (**Figure S6C**) and between PepCan non-recurrence and PepCan recurrence groups (**Figure S6D**) prior to vaccination. We also examined differentially abundant taxa between these groups. Furthermore, alpha diversity was compared and differentially abundant taxa were examined before (at visit 1) and after (at visit 8) vaccination. No significant results were found. Analogous results of amplicon-based rRNA sequencing are shown in **Figure S7**.

### Oral wash

The compositions of oral wash bacteria at phylum and species are level are shown in **Figure S7E** and **Figure S7F** for amplicon-based rRNA sequencing. The comparisons of alphas diversity between PepCan and placebo groups (**Figure S7G**) and between PepCan non-recurrence and PepCan recurrence (**Figure S7H**) are also shown, but there were no significant comparisons.

## DISCUSSION

To our knowledge this is the first study which used a candidate HPV therapeutic vaccine to reduce recurrence of HNC cancer. At present, there are two other clinical trials of HPV therapeutic vaccine trials treating HNC patients that require participants to have no evidence of disease. However, these studies do not mention a goal of prevention of recurrence. These studies utilized mRNA (NCT03418480) or dendritic cell targeting (NCT06007092). Of these, the mRNA study includes an arm for recurrent disease. Likewise, there are several other types of HPV therapeutic vaccines in clinical trials, but their focus is on the treatment of existing HPV or HNC malignancy. There are DNA-based (NCT03162224),^42^ mRNA-based (NCT04534205), cell based (NCT04084951),^43^ viral vector-based (NCT04180215), peptide-based (NCT02426892),^44^ and bacterial vector-based (NCT02002182).

Previous studies which used PepCan to treat CINs demonstrated safety.^18–20^ The current study also showed that PepCan is safe as there were no serious adverse events reported throughout the study. However, there were two DLT events in this study which were grade 2 and 3 allergic reactions, but no DLTs were reported in the previous studies.^18–20^ Previous studies administered four PepCan injections while the current study administered seven. Both DLT events occurred at the 6^th^ injection. Therefore, larger number of injections may predispose to having allergic reactions. The most common injection-related AE was injection site reaction which was significantly more frequent in the PepCan group compared to the placebo group (*p*<0.0001).

Unexpectedly, non-recurrence rate was higher in the placebo group (4/5 or 80% for ITT and PP) compared to the PepCan group (5/11 or 45% for ITT and 5 of 9 or 56% for PP). Some differences in the baseline characteristics such as most patients in the placebo group having T2 disease may have contributed to the unexpected outcome; however, this study is inconclusive because only a small number of patients were enrolled. The expected outcome of the study used for the power calculation (see Methods) was to reduce the recurrence rate from 80%^4,5^ to 44% in the PepCan group. This is very close to the ITT non-recurrence rate of 45%.

Comparison of pre-vaccination T cell responses to HPV 16 E6 showed no difference between the PepCan and placebo groups. Given that most patients did not have HPV-positive HNC, the pre-vaccination HPV 16 E6 responses may be responses to infection to anogenital sites. New post-vaccine responses to HPV 16 E6 were significantly higher in the PepCan non-recurrence group compared to the PepCan recurrence group (*p*=0.05 for ITT and *p*=0.02 for PP) suggesting that the ability to mount T cell responses may be related to clinical efficacy.

There were no differences in the pre-vaccination levels of Th1, Th2, and Treg cells between the PepCan and placebo group (**Figures 2A and S3A**). However, Th1 cells were significantly higher in the PepCan non-recurrence group compared to the recurrence group (**Figures 2B and S2B**, *p*=0.01 for ITT and *p*=0.04 for PP). Therefore, Th1 polarization appears to be associated with vaccine response. This make sense in that *Candida* is known to promote IL-12 production,^6^ and an earlier study using PepCan to treat CIN demonstrated increase in Th1 cells after vaccination.^19^

Cytokine analysis showed significant difference for IL-5 and MCP1-MCAF. The IL-5 levels were found to be significantly higher in the placebo group compared to the PepCan and for the PepCan non-recurrence group compared to the PepCan recurrence group suggesting better clinical outcomes with elevated IL-5 levels (**Figure S5**). The biological significance remains uncertain as the significant increase in IL-5 was found only for the ITT and not PP population. A lower level in the PepCan group would have made more sense as IL-5 is associated with Th2 cells while PepCan is associated with Th1 cells.^45,46^ MCP1-MCAF was higher in placebo compared to PepCan at visit 1 for both the ITT and the PP populations. While not statistically significant, the placebo group showed better outcomes than the PepCan group. This may be explained by the role MCP1 plays in the chemotaxis of memory lymphocytes.^47^

We examined differentially abundant taxa and alpha diversity of the stool samples since it is well know that it influences outcomes of immunotherapy.^36–41^ No significant results were found. Analogous analyses using the oral wash samples did not yield any significant results either likely due to small number of patients enrolled.

The limitation of this study was the small number of patients that were enrolled, and fewer blood, stool, and oral wash samples collected from the recurrence group compared to the non-recurrence group. This was because patients with recurrence completed and exited the study at the time of recurrence and did not necessarily finish all planned study visits.

As the Phase II study of comparing the efficacy of PepCan and *Candida* in regressing CIN unexpectedly showed better outcome with *Candida* (NCT02481414),^20^ we have initiated a new clinical trial enrolling HNC patients with no evidence of disease after standard therapies using *Candida* to reduce recurrence (NCT05952934).

In conclusion, this study demonstrated safety of PepCan although DLT can occur when more injections were given compared to earlier studies.^18–20^ Efficacy could not be assessed given the small number of patients enrolled. New T cell response to HPV 16 E6 protein was associated with non-recurrence group in comparison to recurrence group, and circulating Th1 cells were higher in the recurrence group (**Figure 2B** and **Figure S3B).**

## CONTRIBUTIONS

MN and OA conceptualized the study; EB, OA, TE, GB, JC, RG, AJ, AK, YCWL, KM, MM, JS, EV, MW, and MN participated in data acquisition; EB, TE, MB, ND, JLF, AK, OL, KM, IN, MR, and DU performed data analysis; EB and MN drafted the original version of the manuscript, and all authors edited drafts and approved the final version of the manuscript.

## ACKNOWLEDGEMENTS

The authors would like to thank the patients for participating in this study. The authors would also like to thank Hanah Coleman, Benjamin Lieblong, and Sumit Shah for their technical expertise, James Allred, Nicole Chapman, Angela Trammell, and Amy Jones for their clinical trial expertise, Jennifer Roberts and Amy Crisp for their pharmaceutical expertise, Sorena Lo, James Holley, Madison Trujillo, Larry Parker, Laura Adkins, and Brenda Gannon for their regulatory expertise, and Patricia Gminski and Traci Ireland for their administrative expertise

## DATA AVAILABILITY STATEMENT

The data that support the findings of this study are available from the corresponding author, MN, upon reasonable request.

