## Appendix for "A Randomized Double-Blind Placebo-Controlled Phase I/II Clinical Trial of a Human Papillomavirus Therapeutic Vaccine, PepCan, for Reducing Head and Neck Cancer Recurrence"

### TABLE OF CONTENTS

Supplementary Table 3
Table S1. All adverse events regardless of injection-relatedness

Supplementary Figures 10
Figure S1. Screening visit questionnaire
Figure S2. Representative ELISPOT assay results
Figure S3. FACS analyses of peripheral immune cells per-protocol (PP)
Figure S4. TCR repertoire analyses
Figure S5. Cytokine analyses
Figure S6. Microbiome analyses using metagenomics
Figure S7. Microbiome analyses using amplicon-based rRNA sequencing

Supplementary References 26

#### SUPPLEMENTARY TABLE

##### TABLE S1A. All adverse events regardless of injection-relatedness

| All Adverse Events at the Event Level by Experimental Group and Grade | | CTCAE v5 AE Severity Scale Grade | | | | | | | | | | | |
|---|---|---|---|---|---|---|---|---|---|---|---|---|---|
| | | PepCan | | | | | | Placebo | | | | | |
| **CTCAE v5 System Organ Class** | **CTCAE v5 AE Term** | **1** | **2** | **3** | **4** | **5** | **Total** | **1** | **2** | **3** | **4** | **5** | **Total** |
| Blood and Lymphatic System Disorders [33][u] | Anemia [30][u] | 13[u] | 3[u] | 2[u] | - | - | **18[u]** | 9[u] | 3[u] | - | - | - | **12[u]** |
| | Leukocytosis [1][u] | - | - | - | - | - | **-** | - | - | 1[u] | - | - | **1[u]** |
| | Blood and lymphatic system disorders - Other, specify [1][u] | 1[u] | - | - | - | - | **1[u]** | - | - | - | - | - | **-** |
| Cardiac Disorders [5][u] | Atrial fibrillation [2][u] | - | 1[u] | - | - | - | **1[u]** | - | 1[u] | - | - | - | **1[u]** |
| | Chest pain - cardiac [2][u] | 1[u] | - | 1[u] | - | - | **2[u]** | - | - | - | - | - | **-** |
| | Heart failure [1][u] | - | 1[u] | - | - | - | **1[u]** | - | - | - | - | - | **-** |
| Ear and Labyrinth Disorders [19][u] | Ear pain [3][u] | 3[u] | - | - | - | - | **3[u]** | - | - | - | - | - | **-** |
| | Hearing impaired [6][u] | 3[u] | 1[u] | 1[u] | - | - | **5[u]** | 1[u] | - | - | - | - | **1[u]** |
| | Tinnitus [8][u] | 3[u] | 2[u] | - | - | - | **5[u]** | 3[u] | - | - | - | - | **3[u]** |
| | Vertigo [1][u] | 1[u] | - | - | - | - | **1[u]** | - | - | - | - | - | **-** |
| | Ear and labyrinth disorders - Other, specify [1][u] | 1[u] | - | - | - | - | **1[u]** | - | - | - | - | - | **-** |
| Endocrine Disorders [16][u] | Hyperthyroidism [2][u] | - | 1[u] | - | - | - | **1[u]** | - | 1[u] | - | - | - | **1[u]** |
| | Hypothyroidism [13][u] | 4[u] | 4[u] | - | - | - | **8[u]** | 1[u] | 4[u] | - | - | - | **5[u]** |
| | Endocrine disorders - Other, specify [1][u] | - | - | - | - | - | **-** | 1[u] | - | - | - | - | **1[u]** |
| Eye Disorders [4][u] | Blurred vision [1][u] | - | - | - | - | - | **-** | 1[u] | - | - | - | - | **1[u]** |

| | | | | | | | | | | | | | |
|---|---|---|---|---|---|---|---|---|---|---|---|---|---|
| | Dry eye [1]$^{u}$ | - | - | - | - | - | **-** | $1^{u}$ | - | - | - | - | **$1^{u}$** |
| | Scleral disorder [2]$^{u}$ | $2^{u}$ | - | - | - | - | **$2^{u}$** | - | - | - | - | - | **-** |
| Gastrointestinal Disorders [113]$^{u, 7\ SAE}$ | Abdominal pain [1]$^{u}$ | - | $1^{u}$ | - | - | - | **$1^{u}$** | - | - | - | - | - | **-** |
| | Constipation [9]$^{u, 1\ SAE}$ | $3^{u}$ | $1^{u}$ | - | - | - | **$4^{u}$** | $1^{u}$ | $3^{u}$ | - | $1^{u, SAE}$ | - | **$5^{u, 1\ SAE}$** |
| | Diarrhea [7]$^{u, 2\ SAE}$ | $4^{u}$ | - | $2^{u, SAE}$ | - | - | **$6^{u, 2\ SAE}$** | $1^{u}$ | - | - | - | - | **$1^{u}$** |
| | Dry mouth [24]$^{u}$ | $10^{u}$ | $6^{u}$ | $1^{u}$ | - | - | **$17^{u}$** | $6^{u}$ | $1^{u}$ | - | - | - | **$7^{u}$** |
| | Dysphagia [24]$^{u}$ | $4^{u}$ | $7^{u}$ | $4^{u}$ | - | - | **$15^{u}$** | $3^{u}$ | $4^{u}$ | $2^{u}$ | - | - | **$9^{u}$** |
| | Enterocolitis [1]$^{u}$ | - | - | - | - | - | **-** | - | - | $1^{u}$ | - | - | **$1^{u}$** |
| | Esophageal stenosis [1]$^{u}$ | - | - | - | - | - | **-** | - | $1^{u}$ | - | - | - | **$1^{u}$** |
| | Esophagitis [2]$^{u}$ | - | - | - | - | - | **-** | - | - | $2^{u}$ | - | - | **$2^{u}$** |
| | Gastroesophageal reflux disease [7]$^{u}$ | $2^{u}$ | $2^{u}$ | - | - | - | **$4^{u}$** | $1^{u}$ | $2^{u}$ | - | - | - | **$3^{u}$** |
| | Gastroparesis [1]$^{u}$ | - | $1^{u}$ | - | - | - | **$1^{u}$** | - | - | - | - | - | **-** |
| | Mucositis oral [5]$^{u}$ | - | - | - | - | - | **-** | $1^{u}$ | $3^{u}$ | $1^{u}$ | - | - | **$5^{u}$** |
| | Nausea [4]$^{u}$ | $3^{u}$ | - | - | - | - | **$3^{u}$** | $1^{u}$ | - | - | - | - | **$1^{u}$** |
| | Oral pain [3]$^{u}$ | $1^{u}$ | - | - | - | - | **$1^{u}$** | $2^{u}$ | - | - | - | - | **$2^{u}$** |
| | Pancreatitis [1]$^{u}$ | - | $1^{u}$ | - | - | - | **$1^{u}$** | - | - | - | - | - | **-** |
| | Upper gastrointestinal hemorrhage [1]$^{u, SAE}$ | - | - | - | - | - | **-** | - | - | $1^{u, SAE}$ | - | - | **$1^{u, SAE}$** |
| | Vomiting [5]$^{u, 1\ SAE}$ | $3^{u}$ | - | $1^{u, SAE}$ | - | - | **$4^{u, 1\ SAE}$** | - | - | $1^{u}$ | - | - | **$1^{u}$** |
| | Gastrointestinal disorders - Other, specify [17]$^{u, 2\ SAE}$ | $5^{u}$ | $3^{u}$ | $2^{u, SAE}$ | - | - | **$10^{u, 2\ SAE}$** | $3^{u}$ | $4^{u}$ | - | - | - | **$7^{u}$** |
| General Disorders and Site Administration Conditions [53]$^{u29, 1\ SAE}$ | Chills [1]$^{u}$ | $1^{u}$ | - | - | - | - | **$1^{u}$** | - | - | - | - | - | **-** |
| | Edema face [4]$^{u}$ | $2^{u}$ | - | - | - | - | **$2^{u}$** | $2^{u}$ | - | - | - | - | **$2^{u}$** |
| | Facial pain [2]$^{u}$ | $1^{u}$ | $1^{u}$ | - | - | - | **$2^{u}$** | - | - | - | - | - | **-** |
| | Fatigue [13]$^{u12}$ | $6^{u}$ | $2^{u}$ | - | - | - | **$8^{u}$** | $5^{u4}$ | - | - | - | - | **$5^{u4}$** |

| | | | | | | | | | | | | | |
|---|---|---|---|---|---|---|---|---|---|---|---|---|---|
| | Fever [1]$^{u}$ | 1$^{u}$ | - | - | - | - | **1$^{u}$** | - | - | - | - | - | **-** |
| | Gait disturbance [2]$^{u, 1 SAE}$ | - | 2$^{u,}$ 1 SAE | - | - | - | **2$^{u, 1}$ SAE** | - | - | - | - | - | **-** |
| | Injection site reaction [23] | 20 | 3 | - | - | - | **23** | - | - | - | - | - | **-** |
| | Malaise [1]$^{u}$ | 1$^{u}$ | - | - | - | - | **1$^{u}$** | - | - | - | - | - | **-** |
| | Neck edema [4]$^{u}$ | 3$^{u}$ | - | - | - | - | **3$^{u}$** | 1$^{u}$ | - | - | - | - | **1$^{u}$** |
| | Pain [1]$^{u}$ | 1$^{u}$ | - | - | - | - | **1$^{u}$** | - | - | - | - | - | **-** |
| | General disorders and administration site conditions - Other, specify [1]$^{u}$ | 1$^{u}$ | - | - | - | - | **1$^{u}$** | - | - | - | - | - | **-** |
| Immune System Disorders [3]$^{u1}$ | Allergic reaction [3]$^{u1}$ | - | 1 | 2$^{u}$ 1 | - | - | **3$^{u1}$** | - | - | - | - | - | **-** |
| Infections and Infestations [9]$^{u, 1 SAE}$ | Bacteremia [1]$^{u,}$ SAE | - | 1$^{u,}$ SAE | - | - | - | **1$^{u,}$ SAE** | - | - | - | - | - | **-** |
| | Bone infection [1]$^{u}$ | - | - | 1$^{u}$ | - | - | **1$^{u}$** | - | - | - | - | - | **-** |
| | Shingles [1]$^{u}$ | - | 1$^{u}$ | - | - | - | **1$^{u}$** | - | - | - | - | - | **-** |
| | Sinusitis [1]$^{u}$ | - | - | - | - | - | **-** | - | 1$^{u}$ | - | - | - | **1$^{u}$** |
| | Skin infection [1]$^{u}$ | - | 1$^{u}$ | - | - | - | **1$^{u}$** | - | - | - | - | - | **-** |
| | Thrush [1]$^{u}$ | - | - | - | - | - | **-** | - | 1$^{u}$ | - | - | - | **1$^{u}$** |
| | Upper respiratory infection [2]$^{u}$ | - | 1$^{u}$ | - | - | - | **1$^{u}$** | - | 1$^{u}$ | - | - | - | **1$^{u}$** |
| | Infections and infestations - Other, specify [1]$^{u}$ | - | 1$^{u}$ | - | - | - | **1$^{u}$** | - | - | - | - | - | **-** |
| Injury, Poisoning and Procedural Complications [9]$^{u4}$ | Bruising [1]$^{u}$ | - | - | - | - | - | **-** | 1$^{u}$ | - | - | - | - | **1$^{u}$** |
| | Dermatitis radiation [3]$^{u}$ | 1$^{u}$ | 1$^{u}$ | - | - | - | **2$^{u}$** | - | 1$^{u}$ | - | - | - | **1$^{u}$** |
| | Vaccination complication [5] | 3 | 1 | - | - | - | **4** | 1 | - | - | - | - | **1** |

| | | | | | | | | | | | | | |
|---|---|---|---|---|---|---|---|---|---|---|---|---|---|
| Investigations [164][u, 1 SAE] | Alanine aminotransferase increased [1][u] | - | - | - | - | - | **-** | 1[u] | - | - | - | - | **1[u]** |
| | Alkaline phosphatase increased [7][u] | 7[u] | - | - | - | - | **7[u]** | - | - | - | - | - | **-** |
| | Aspartate aminotransferase increased [2][u] | 1[u] | - | - | - | - | **1[u]** | 1[u] | - | - | - | - | **1[u]** |
| | Blood antidiuretic hormone abnormal [1][u] | - | - | - | - | - | **-** | 1[u] | - | - | - | - | **1[u]** |
| | Blood bilirubin increased [3][u] | 2[u] | 1[u] | - | - | - | **3[u]** | - | - | - | - | - | **-** |
| | Cholesterol high [1][u] | 1[u] | - | - | - | - | **1[u]** | - | - | - | - | - | **-** |
| | Creatinine increased [6][u] | 3[u] | - | - | - | - | **3[u]** | 3[u] | - | - | - | - | **3[u]** |
| | Lymphocyte count decreased [53][u] | 14[u] | 16[u] | 4[u] | - | - | **34[u]** | 6[u] | 9[u] | 4[u] | - | - | **19[u]** |
| | Neutrophil count decreased [4][u] | 1[u] | 2[u] | - | - | - | **3[u]** | - | 1[u] | - | - | - | **1[u]** |
| | Platelet count decreased [4][u] | 4[u] | - | - | - | - | **4[u]** | - | - | - | - | - | **-** |
| | Weight loss [11][u, 1 SAE] | 5[u] | 3[u] | 2[u, 1 SAE] | - | - | **10[u, 1 SAE]** | 1[u] | - | - | - | - | **1[u]** |
| | White blood cell decreased [20][u] | 6[u] | 4[u] | - | - | - | **10[u]** | 8[u] | 2[u] | - | - | - | **10[u]** |
| | Investigations - Other, specify [46][u] | 31[u] | 2[u] | - | - | - | **33[u]** | 12[u] | 1[u] | - | - | - | **13[u]** |
| Metabolism and Nutrition Disorders [95][u94, 1 SAE] | Anorexia [10][u] | 1[u] | 3[u] | 2[u] | - | - | **6[u]** | 2[u] | 1[u] | 1[u] | - | - | **4[u]** |
| | Hypercalcemia [4][u] | 4[u] | - | - | - | - | **4[u]** | - | - | - | - | - | **-** |
| | Hyperglycemia [13][u] | 8[u] | 1[u] | - | - | - | **9[u]** | 3[u] | 1[u] | - | - | - | **4[u]** |
| | Hyperkalemia [1][u] | - | - | - | - | - | **-** | 1[u] | - | - | - | - | **1[u]** |
| | Hyperlipidemia [2][u] | - | 1[u] | - | - | - | **1[u]** | - | 1[u] | - | - | - | **1[u]** |
| | Hypoalbuminemia [13][u] | 3[u] | 2[u] | - | - | - | **5[u]** | 8[u] | - | - | - | - | **8[u]** |
| | Hypocalcemia [16][u, 1 SAE] | 8[u] | 4[u] | 2[u] | 1[u, SAE] | - | **15[u, 1 SAE]** | 1[u] | - | - | - | - | **1[u]** |

| | | | | | | | | | | | | | |
|---|---|---|---|---|---|---|---|---|---|---|---|---|---|
| | Hypoglycemia [1]$^{u}$ | 1$^{u}$ | - | - | - | - | **1$^{u}$** | - | - | - | - | - | **-** |
| | Hypokalemia [2]$^{u1}$ | 2$^{u1}$ | - | - | - | - | **2$^{u1}$** | - | - | - | - | - | **-** |
| | Hypomagnesemia [1]$^{u}$ | 1$^{u}$ | - | - | - | - | **1$^{u}$** | - | - | - | - | - | **-** |
| | Hyponatremia [20]$^{u}$ | 9$^{u}$ | - | 1$^{u}$ | - | - | **10$^{u}$** | 8$^{u}$ | - | 1$^{u}$ | 1$^{u}$ | - | **10$^{u}$** |
| | Hypophosphatemia [2]$^{u}$ | 1$^{u}$ | - | - | - | - | **1$^{u}$** | 1$^{u}$ | - | - | - | - | **1$^{u}$** |
| | Metabolism and nutrition disorders - Other, specify [4]$^{u}$ | - | 3$^{u}$ | - | - | - | **3$^{u}$** | 1$^{u}$ | - | - | - | - | **1$^{u}$** |
| Musculoskeletal and Connective Tissue Disorders [22]$^{u, 2 SAE}$ | Arthritis [2]$^{u}$ | 1$^{u}$ | 1$^{u}$ | - | - | - | **2$^{u}$** | - | - | - | - | - | **-** |
| | Back pain [7]$^{u}$ | 2$^{u}$ | 2$^{u}$ | - | - | - | **4$^{u}$** | 2$^{u}$ | 1$^{u}$ | - | - | - | **3$^{u}$** |
| | Generalized muscle weakness [1]$^{u}$ | - | 1$^{u}$ | - | - | - | **1$^{u}$** | - | - | - | - | - | **-** |
| | Muscle weakness lower limb [1]$^{u}$ | 1$^{u}$ | - | - | - | - | **1$^{u}$** | - | - | - | - | - | **-** |
| | Muscle weakness upper limb [1]$^{u}$ | 1$^{u}$ | - | - | - | - | **1$^{u}$** | - | - | - | - | - | **-** |
| | Neck pain [1]$^{u}$ | 1$^{u}$ | - | - | - | - | **1$^{u}$** | - | - | - | - | - | **-** |
| | Osteonecrosis of jaw [5]$^{u, 2 SAE}$ | - | 3$^{u,}$ 1 SAE | 2$^{u,}$ 1 SAE | - | - | **5$^{u, 2}$ SAE** | - | - | - | - | - | **-** |
| | Osteoporosis [1]$^{u}$ | - | - | - | - | - | **-** | 1$^{u}$ | - | - | - | - | **1$^{u}$** |
| | Pain in extremity [2]$^{u}$ | - | 2$^{u}$ | - | - | - | **2$^{u}$** | - | - | - | - | - | **-** |
| | Musculoskeletal and connective tissue disorder - Other, specify [1]$^{u}$ | 1$^{u}$ | - | - | - | - | **1$^{u}$** | - | - | - | - | - | **-** |
| Neoplasms benign, malignant and unspecified (incl cysts and polyps) [5]$^{u, 1 SAE}$ | Tumor pain [3]$^{u}$ | 1$^{u}$ | 2$^{u}$ | - | - | - | **3$^{u}$** | - | - | - | - | - | **-** |
| | Neoplasms benign, malignant and unspecified (incl cysts and polyps) - Other, specify [2]$^{u, 1 SAE}$ | - | 1$^{u}$ | - | - | 1$^{u,}$ SAE | **2$^{u, 1}$ SAE** | - | - | - | - | - | **-** |
| Nervous System Disorders [34]$^{u31}$ | Dizziness [4]$^{u3}$ | 3$^{u2}$ | - | - | - | - | **3$^{u2}$** | 1$^{u}$ | - | - | - | - | **1$^{u}$** |
| | Dysgeusia [13]$^{u}$ | 6$^{u}$ | 4$^{u}$ | - | - | - | **10$^{u}$** | 3$^{u}$ | - | - | - | - | **3$^{u}$** |

| | | | | | | | | | | | | | |
|---|---|---|---|---|---|---|---|---|---|---|---|---|---|
| | Dysphasia [1]$^{u}$ | 1$^{u}$ | - | - | - | - | **1$^{u}$** | - | - | - | - | - | **-** |
| | Headache [7]$^{u5}$ | 3$^{u1}$ | 1$^{u}$ | 1$^{u}$ | - | - | **5$^{u3}$** | - | 2$^{u}$ | - | - | - | **2$^{u}$** |
| | Memory impairment [1]$^{u}$ | 1$^{u}$ | - | - | - | - | **1$^{u}$** | - | - | - | - | - | **-** |
| | Neuralgia [1]$^{u}$ | - | - | - | - | - | **-** | 1$^{u}$ | - | - | - | - | **1$^{u}$** |
| | Paresthesia [3]$^{u}$ | 2$^{u}$ | - | - | - | - | **2$^{u}$** | 1$^{u}$ | - | - | - | - | **1$^{u}$** |
| | Peripheral motor neuropathy [1]$^{u}$ | - | - | - | - | - | **-** | 1$^{u}$ | - | - | - | - | **1$^{u}$** |
| | Peripheral sensory neuropathy [2]$^{u}$ | - | 1$^{u}$ | - | - | - | **1$^{u}$** | - | 1$^{u}$ | - | - | - | **1$^{u}$** |
| | Spasticity [1]$^{u}$ | 1$^{u}$ | - | - | - | - | **1$^{u}$** | - | - | - | - | - | **-** |
| Psychiatric Disorders [16]$^{u}$ | Anxiety [5]$^{u}$ | - | 3$^{u}$ | - | - | - | **3$^{u}$** | 1$^{u}$ | 1$^{u}$ | - | - | - | **2$^{u}$** |
| | Depression [7]$^{u}$ | 3$^{u}$ | 1$^{u}$ | - | - | - | **4$^{u}$** | 1$^{u}$ | 2$^{u}$ | - | - | - | **3$^{u}$** |
| | Insomnia [2]$^{u}$ | 1$^{u}$ | - | - | - | - | **1$^{u}$** | - | 1$^{u}$ | - | - | - | **1$^{u}$** |
| | Restlessness [2]$^{u}$ | - | - | - | - | - | **-** | 1$^{u}$ | 1$^{u}$ | - | - | - | **2$^{u}$** |
| Renal and Urinary Disorders [9]$^{u,}$ 1 SAE | Acute kidney injury [1]$^{u}$ | - | - | - | - | - | **-** | - | - | 1$^{u}$ | - | - | **1$^{u}$** |
| | Chronic kidney disease [2]$^{u}$ | - | 1$^{u}$ | - | - | - | **1$^{u}$** | - | 1$^{u}$ | - | - | - | **1$^{u}$** |
| | Dysuria [1]$^{u}$ | 1$^{u}$ | - | - | - | - | **1$^{u}$** | - | - | - | - | - | **-** |
| | Proteinuria [1]$^{u}$ | 1$^{u}$ | - | - | - | - | **1$^{u}$** | - | - | - | - | - | **-** |
| | Urinary frequency [2]$^{u}$ | - | - | - | - | - | **-** | 1$^{u}$ | 1$^{u}$ | - | - | - | **2$^{u}$** |
| | Urinary tract obstruction [1]$^{u,}$ SAE | - | - | - | - | - | **-** | - | - | 1$^{u,}$ SAE | - | - | **1$^{u,}$ SAE** |
| | Urinary urgency [1]$^{u}$ | - | - | - | - | - | **-** | - | 1$^{u}$ | - | - | - | **1$^{u}$** |
| Reproductive System and Breast Disorders [4]$^{u}$ | Erectile dysfunction [1]$^{u}$ | - | - | - | - | - | **-** | 1$^{u}$ | - | - | - | - | **1$^{u}$** |
| | Reproductive system and breast disorders - Other, specify [3]$^{u}$ | - | 2$^{u}$ | - | - | - | **2$^{u}$** | - | 1$^{u}$ | - | - | - | **1$^{u}$** |
| Respiratory, Thoracic and | Allergic rhinitis [1]$^{u}$ | 1$^{u}$ | - | - | - | - | **1$^{u}$** | - | - | - | - | - | **-** |

| | | | | | | | | | | | | | |
|---|---|---|---|---|---|---|---|---|---|---|---|---|---|
| Mediastinal Disorders [31][u] | Cough [5][u] | 2[u] | 1[u] | - | - | - | **3[u]** | 1[u] | 1[u] | - | - | - | **2[u]** |
| | Dyspnea [4][u] | 2[u] | 1[u] | - | - | - | **3[u]** | 1[u] | - | - | - | - | **1[u]** |
| | Epistaxis [1][u] | 1[u] | - | - | - | - | **1[u]** | - | - | - | - | - | **-** |
| | Nasal congestion [1][u] | 1[u] | - | - | - | - | **1[u]** | - | - | - | - | - | **-** |
| | Oropharyngeal pain [1][u] | - | - | - | - | - | **-** | 1[u] | - | - | - | - | **1[u]** |
| | Pharyngolarynge al pain [1][u] | - | 1[u] | - | - | - | **1[u]** | - | - | - | - | - | **-** |
| | Postnasal drip [1][u] | 1[u] | - | - | - | - | **1[u]** | - | - | - | - | - | **-** |
| | Rhinorrhea [2][u] | 1[u] | - | - | - | - | **1[u]** | 1[u] | - | - | - | - | **1[u]** |
| | Sore throat [10][u] | 4[u] | - | 1[u] | - | - | **5[u]** | 4[u] | 1[u] | - | - | - | **5[u]** |
| | Wheezing [1][u] | 1[u] | - | - | - | - | **1[u]** | - | - | - | - | - | **-** |
| | Respiratory, thoracic and mediastinal disorders - Other, specify [3][u] | - | 2[u] | - | - | - | **2[u]** | - | 1[u] | - | - | - | **1[u]** |
| Skin and Subcutaneous Tissue Disorders [6][u5] | Dry skin [2][u] | 2[u] | - | - | - | - | **2[u]** | - | - | - | - | - | **-** |
| | Pruritus [2][u1] | - | 1 | - | - | - | **1** | 1[u] | - | - | - | - | **1[u]** |
| | Skin and subcutaneous tissue disorders - Other, specify [2][u] | 2[u] | - | - | - | - | **2[u]** | - | - | - | - | - | **-** |
| Surgical and medical procedures [8][u, 4 SAE] | Surgical and medical procedures - Other, specify [8][u, 4 SAE] | - | 2[u] | 5[u, 3 SAE] | - | - | **7[u, 3 SAE]** | - | - | 1[u, SAE] | - | - | **1[u, SAE]** |
| Vascular Disorders [21][u, 1 SAE] Vascular Disorders [21][u, 1 SAE] | Hematoma [1][u] | 1[u] | - | - | - | - | **1[u]** | - | - | - | - | - | **-** |
| | Hypertension [13][u] | 2[u] | 4[u] | 2[u] | - | - | **8[u]** | 1[u] | 1[u] | 3[u] | - | - | **5[u]** |
| | Hypotension [1][u] | - | 1[u] | - | - | - | **1[u]** | - | - | - | - | - | **-** |
| | Lymphedema [2][u] | 1[u] | - | - | - | - | **1[u]** | 1[u] | - | - | - | - | **1[u]** |
| | Vascular disorders - Other, specify [4][u, 1 SAE] | - | 1[u] | - | - | - | **1[u]** | 1[u] | 1[u] | 1[u, SAE] | - | - | **3[u, 1 SAE]** |

U = unrelated to treatment. SAE = serious adverse event. The highest grade shown for each adverse event.

#### SUPPLEMENTARY FIGURES AND FIGURE LEGENDS

**Figure S1. Screening visit questionnaire.** Patients who consented for the study and showed up to the screening visit (n=18) were given a questionnaire asking about whether HPV vaccine was received, their motivation for participation, employment, education, number of children, number of lifetime sexual partners, types of sexual activity, and use of alcohol, tobacco or other nicotine products. The x-axis represents number of patients.

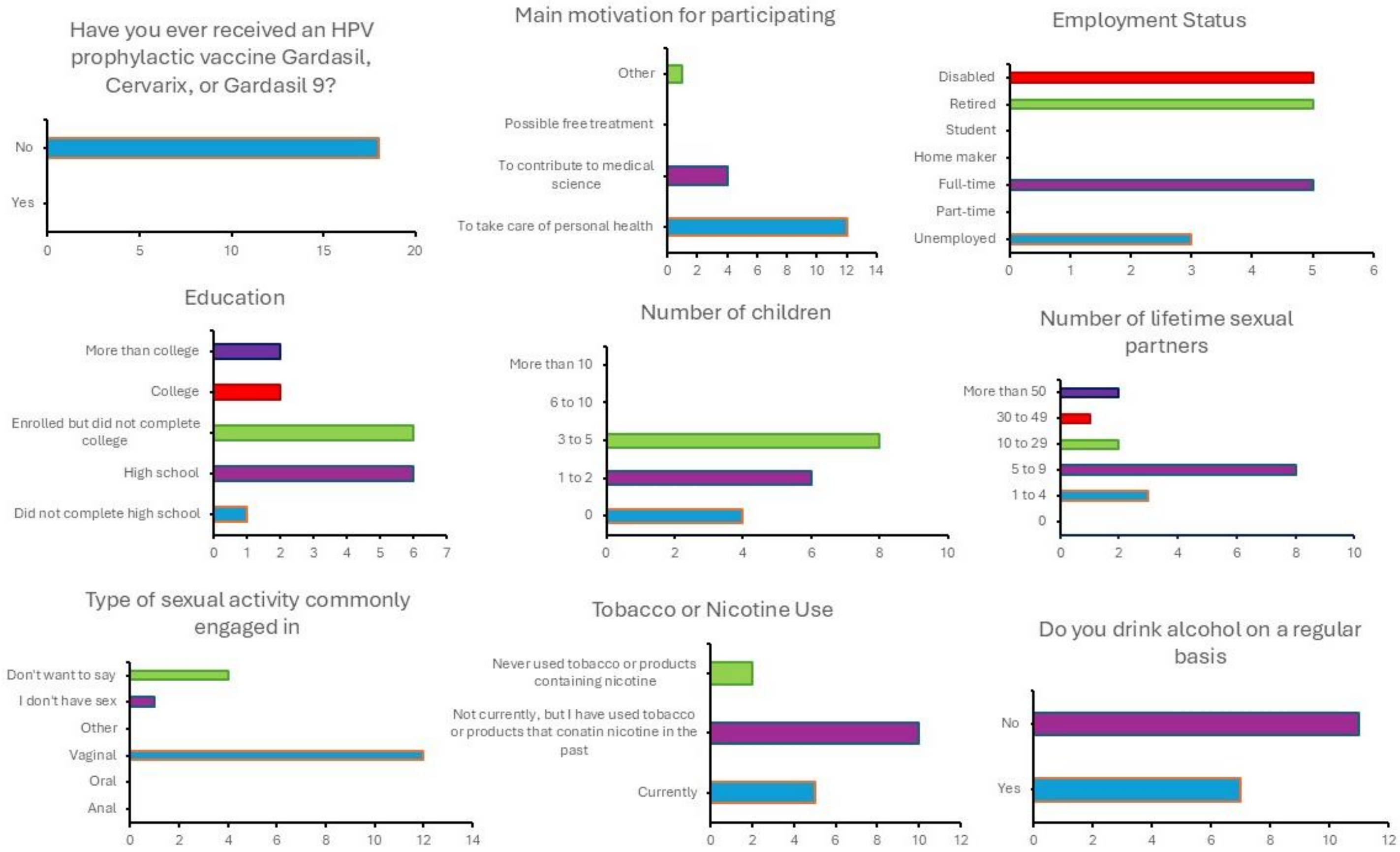

**Figure S2. Representative ELISPOT assay results.** Patient 1 was a PepCan recipient with non-recurrence, and he developed T cell responses to the HPV 16 E6 76-100 region after vaccination. Patient 12 was a PepCan recipient with recurrence, and he did not develop any detectable responses to HPV 16 E6. Positivity indices to peptide pools are shown. Phytohemagglutinin was used as a positive control (not shown). The positivity indices to phytohemagglutinin were 34.2, 61.8, 47.4, and 36 for visits 1, 5, 8 and 9 respectively for patient 1. They were 29.5, 48.6, and 41.5 for visits 1, 5, 8 respectively for patient 12.

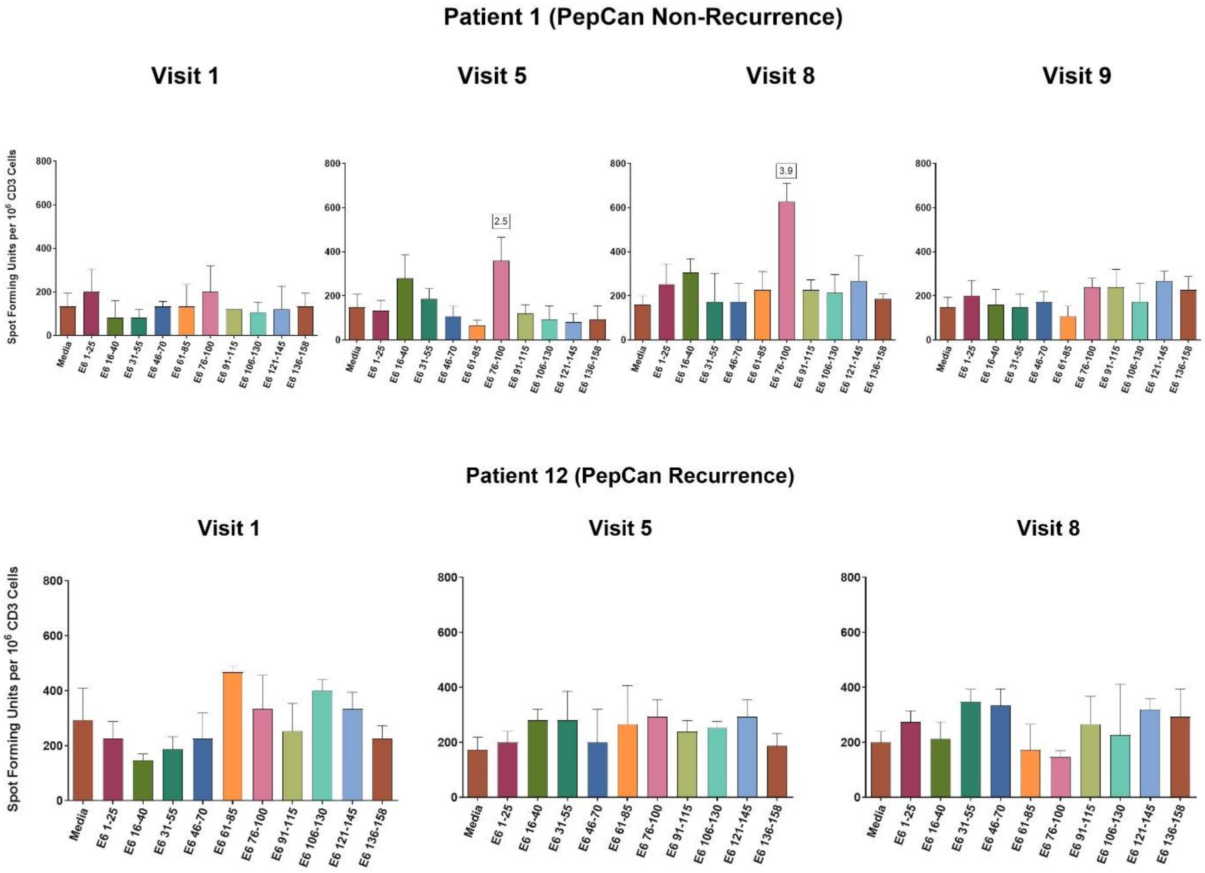

**Figure S3. FACS analyses of peripheral immune cells per-protocol (PP)** Immune cells in peripheral blood were analyzed before and after vaccinations. **A.** Analyses of T-helper type 1 (Th1), T-helper type 2 (Th2), and regulatory T cell (Treg) at visit 1 (pre-vaccination) between the PepCan and the placebo groups. **B**. Analyses of Th1, and Th2, and Treg also at visit 1 between the PepCan non-recurrence group compared to the PepCan recurrence group. **C**. Analyses of Th1, and Th2, and Treg also at visits 1, 5, 8, and 9 between the PepCan and placebo groups. **D**. Analyses of Th1, Th2, and Treg at visits 1, 5, 8, and 9 between non-recurrence PepCan group and recurrence PepCan group.

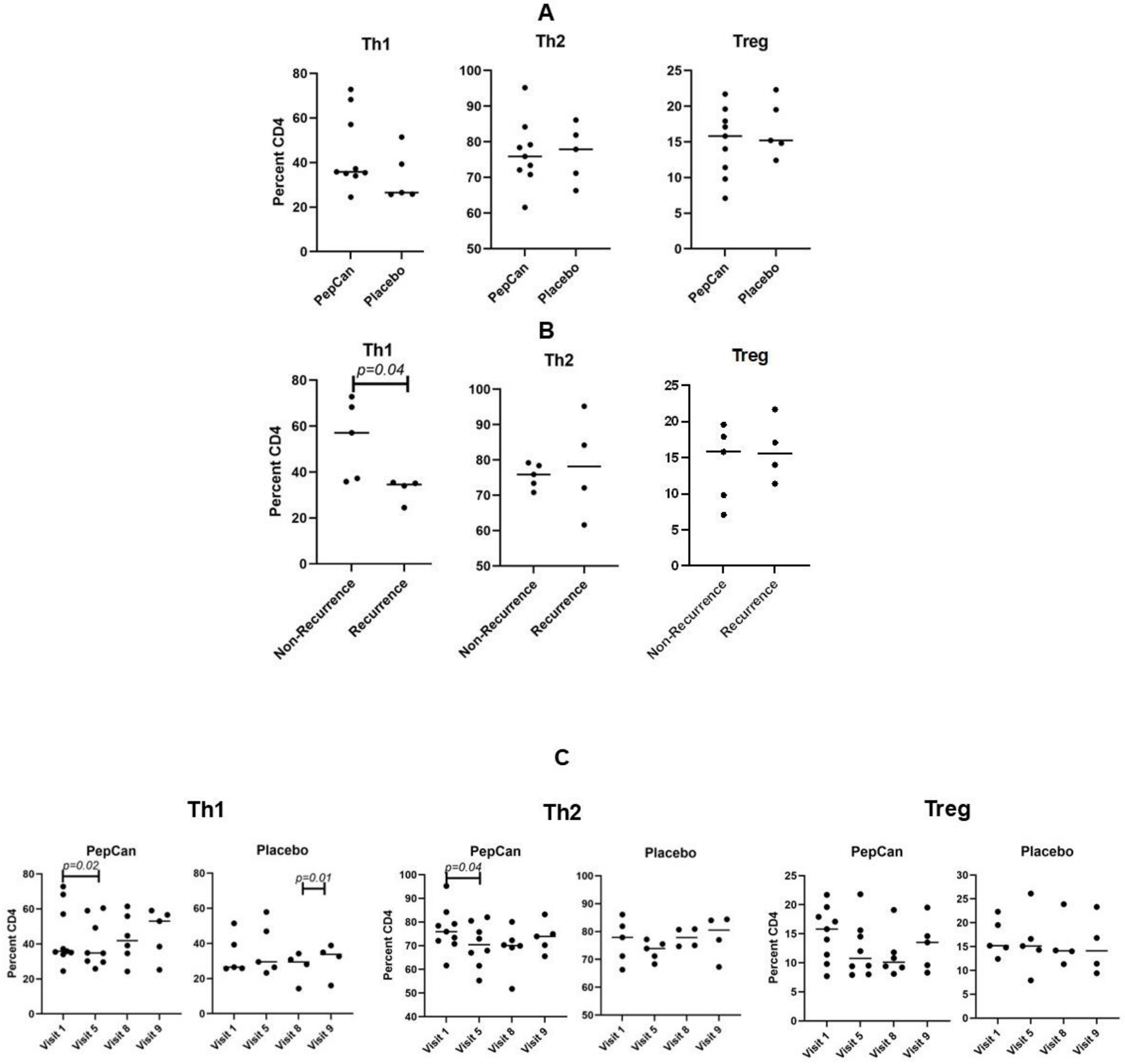

**D**

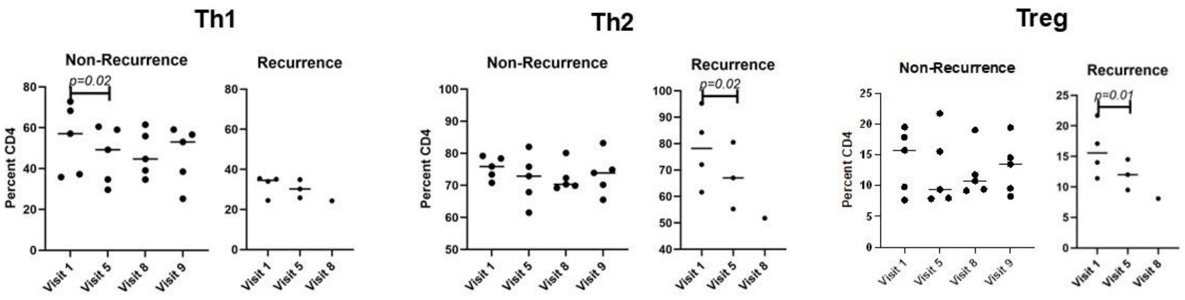
Th1
Non-Recurrence
Recurrence
p=0.02
Percent CD4
Visit 1
Visit 5
Visit 8
Visit 9
Th2
Non-Recurrence
Recurrence
p=0.02
Percent CD4
Treg
Non-Recurrence
Recurrence
p=0.01
Percent CD4

**Figure S4. TCR repertoire analyses** Bulk T cell receptor (TCR) β deep sequencing of peripheral blood mononuclear cells (PBMCs) was performed (n=16)(Adaptive Biotechnologies, Seattle, WA). Putatively vaccine-specific T cells were identified by comparing post-vaccination PBMC sample to the pre-vaccination PBMC sample using a beta-binomial model.[1] **A**. T cell repertoire analysis of the PepCan and placebo groups [intention-to-treat (ITT)] using the numbers of putatively vaccine-specific templates. **B**. T cell repertoire analyses of the PepCan non-recurrence and the PepCan recurrence groups (ITT) using the numbers of putatively vaccine-specific templates. **C**. T cell repertoire analysis of the PepCan and placebo groups (ITT) using the fractions of putatively vaccine-specific T cells among all T cells. **D**. T cell repertoire analyses of the PepCan non-recurrence and the PepCan recurrence groups (ITT) using the fractions of putatively vaccine-specific T cells. **E**. T cell repertoire analysis of the PepCan and placebo groups (PP) using the numbers of putatively vaccine-specific templates. **F**. T cell repertoire analyses of the PepCan non-recurrence and the PepCan recurrence groups (PP) using the numbers of putatively vaccine-specific templates. **G**. T cell repertoire analysis of the PepCan and placebo groups (PP) using the fractions of putatively vaccine-specific T cells among all T cells. **H**. T cell repertoire analyses of the PepCan non-recurrence and the PepCan recurrence groups (PP) using the fractions of putatively vaccine-specific T cells. No significant differences were found for all comparisons.

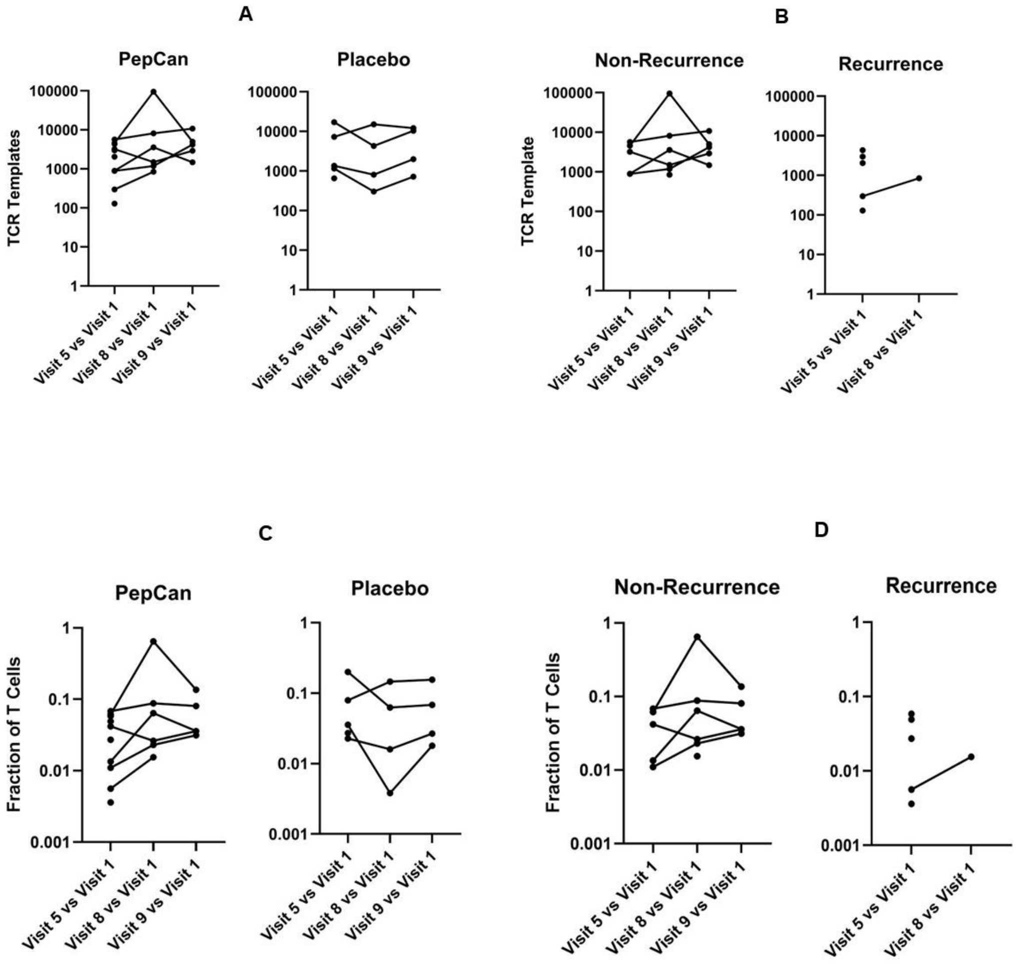

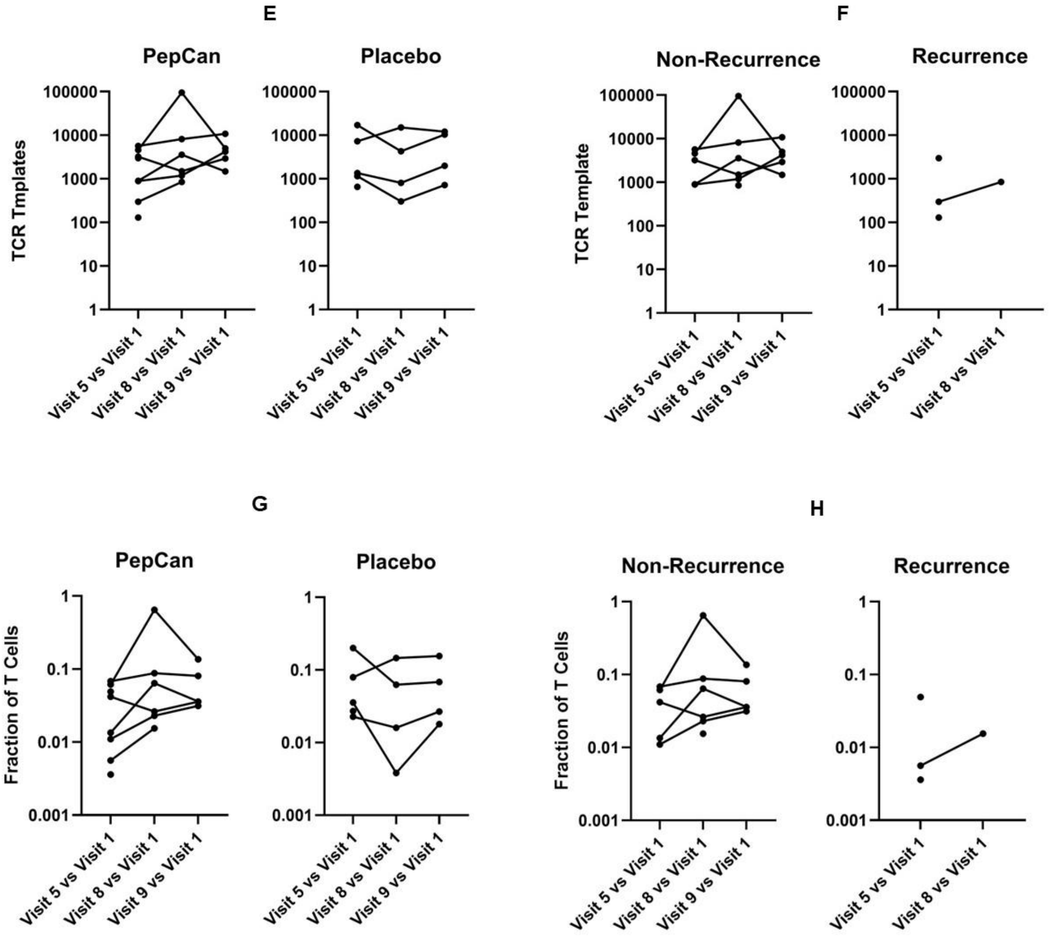
E
PepCan
Placebo
TCR Tmplates
100000
10000
1000
100
10
1
Visit 5 vs Visit 1
Visit 8 vs Visit 1
Visit 9 vs Visit 1
F
Non-Recurrence
Recurrence
TCR Template
G
PepCan
Placebo
Fraction of T Cells
1
0.1
0.01
0.001
H
Non-Recurrence
Recurrence

**Figure S5. Cytokine analyses.** Analyses were performed using a two-level random intercept linear mixed effects model (with cytokines at different visits nested within patients). The 48-plex data (48 biomarkers), inflammation data (30 biomarkers), and TGF-β data (3 biomarkers) were analyzed separately. Two biomarkers from the 48-plex data and ten biomarkers from inflammation data had more than 50% of values undetectable, and were excluded from further analysis. The Shapiro-Wilk test was used to test the deviation from normality assumption for the cytokine measures separately by study visit. An interaction variable for group by time/visit was included in the model to assess whether between-group differences varied across time. In the event that the interaction trended towards statistically significant ($p$-value < 0.10) effect, the between-group effect size was estimated separately at each visit; and longitudinal changes in the biomarkers were assessed separately within each study group. Bonferroni adjusted $p$-value were obtained for post-hoc comparisons. Finally, the false discovery rate (FDR) correction was applied to $p$-values across all the biomarkers included in the analysis. **A**. Significant comparisons from the ITT analyses comparing PepCan and placebo groups are shown. The mean IL5 level was significantly lower in the PepCan group compared to placebo group (Difference: -227.91, 95% CI: -372.88 to -82.95, $p < 0.01$). For MCP1_MCAF and SDF1a, the interaction effect was retained. Treatment effects were assessed at individual visits, and longitudinal trends were examined within each group. At visit 1, the mean MCP1_MCAF level was significantly lower in the PepCan group compared to the placebo group (Difference: -34.38; 95% CI: -56.76 to -12.00; $p < 0.01$). Among placebo-treated patients, the mean SDF1a level was significantly lower at visit 8 (Difference: -785.39; 95% CI: -1246.60 to -324.18; $p < 0.01$) and visit 9 (Difference: -625.51; 95% CI: -1086.72 to -164.30; $p < 0.01$) compared to visit 1. Additionally, a significant decrease in mean BAFF_TNFSF13B was observed among placebo-treated patients at Visit 8 (Difference: -9556.52; 95% CI: -15,903.43 to -3209.61; $p < 0.01$) and at Visit 9 (Difference: -8622.74; 95% CI: -14,969.65 to -2275.83; $p < 0.01$), both compared to visit 1. **B**. Significant comparisons from the ITT analyses comparing PepCan non-recurrence and PepCan recurrence groups are shown. The mean IL5 level was significantly higher among non-recurrent patients compared to recurrent patients (Difference: 215.49, 95% CI: -86.81 to 344.17, $p < 0.01$). For IL1ra, IL12_p40, and BAFF_TNFSF13B the interaction effect was retained. For all three biomarkers, non-recurrent patients had a significantly lower mean levels at visit 5 compared to visit 1 ($p < 0.01$). In recurrent patients, mean IL28A_IFNl2 level was significantly higher at visit 5 compared to visit 1 ($p < 0.01$).

**A**

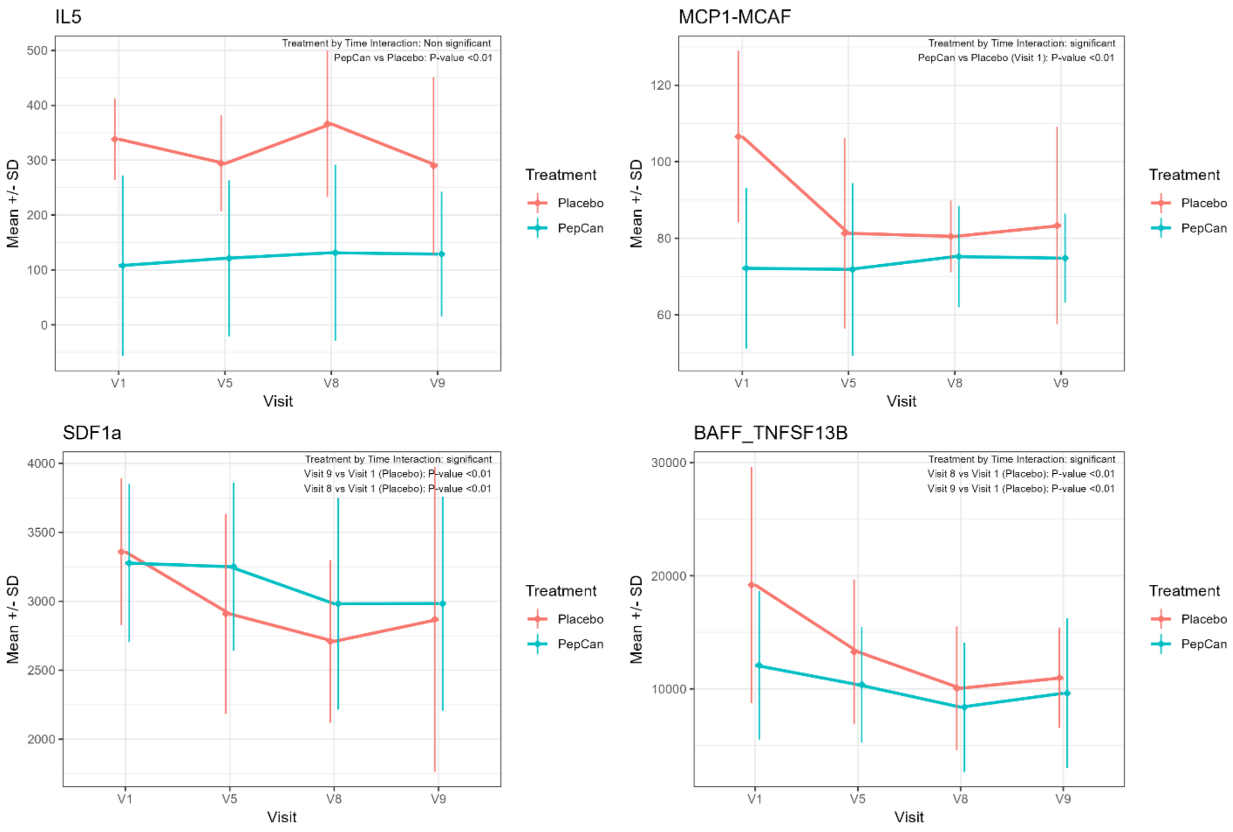
IL5
Treatment by Time Interaction: Non significant
PepCan vs Placebo: P-value <0.01
Mean +/- SD
Visit
V1
V5
V8
V9
Treatment
Placebo
PepCan
MCP1-MCAF
Treatment by Time Interaction: significant
PepCan vs Placebo (Visit 1): P-value <0.01
SDF1a
Treatment by Time Interaction: significant
Visit 9 vs Visit 1 (Placebo): P-value <0.01
Visit 8 vs Visit 1 (Placebo): P-value <0.01
BAFF_TNFSF13B
Treatment by Time Interaction: significant
Visit 8 vs Visit 1 (Placebo): P-value <0.01
Visit 9 vs Visit 1 (Placebo): P-value <0.01

**B**

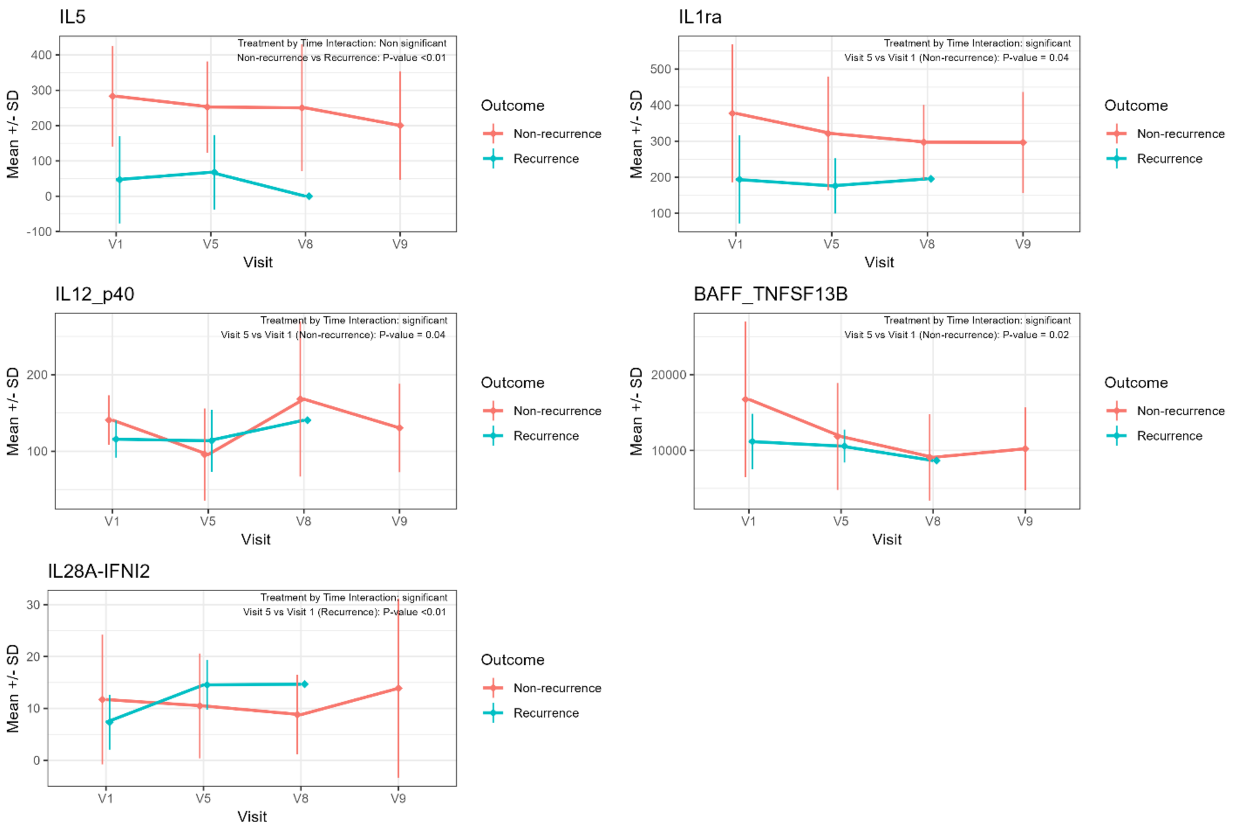
IL5
Treatment by Time Interaction: Non significant
Non-recurrence vs Recurrence: P-value <0.01
Mean +/- SD
Visit
V1
V5
V8
V9
Outcome
Non-recurrence
Recurrence
IL1ra
Treatment by Time Interaction: significant
Visit 5 vs Visit 1 (Non-recurrence): P-value = 0.04
IL12_p40
Treatment by Time Interaction: significant
Visit 5 vs Visit 1 (Non-recurrence): P-value = 0.04
BAFF_TNFSF13B
Treatment by Time Interaction: significant
Visit 5 vs Visit 1 (Non-recurrence): P-value = 0.02
IL28A-IFNl2
Treatment by Time Interaction: significant
Visit 5 vs Visit 1 (Recurrence): P-value <0.01

**Figure S6. Microbiome analyses using metagenomics A**. Phyla in individual stool samples are shown. Each column represents a stool sample from a particular visit of a particular patient. **B**. The 60 highest relative abundant species in individual stool samples are shown. **C**. Comparisons of alpha diversity metrics between PepCan and placebo groups in pre-vaccination (visit 1) stool samples. No significant differences were found. **D**. Comparisons of alpha diversity metrics between PepCan recurrence and PepCan non-recurrence groups in pre-vaccination (visit 1) stool samples. No significant differences were found.

**A**

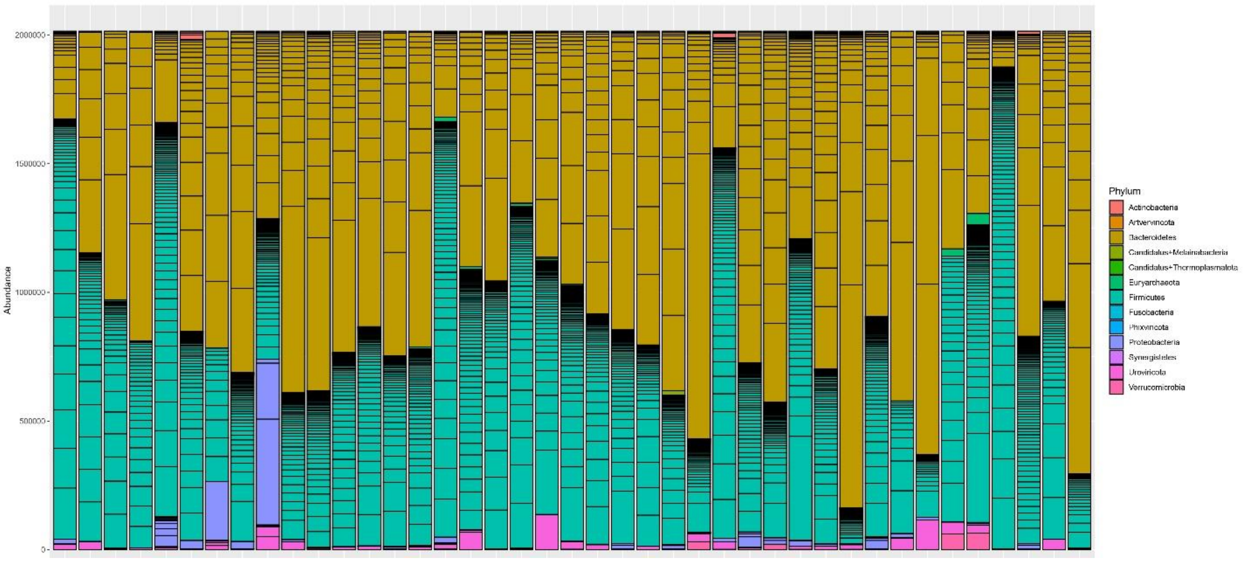


**B**

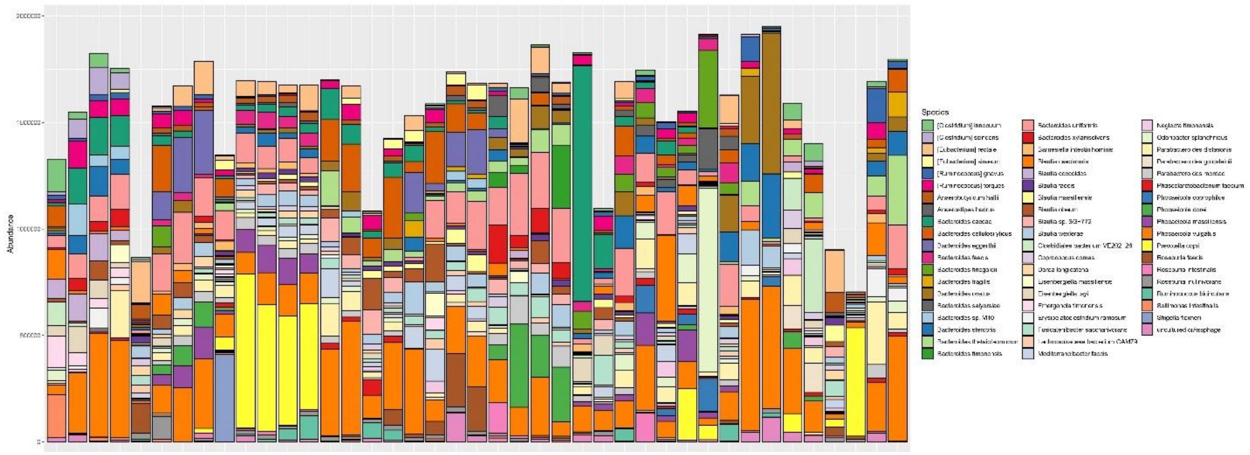

**C**

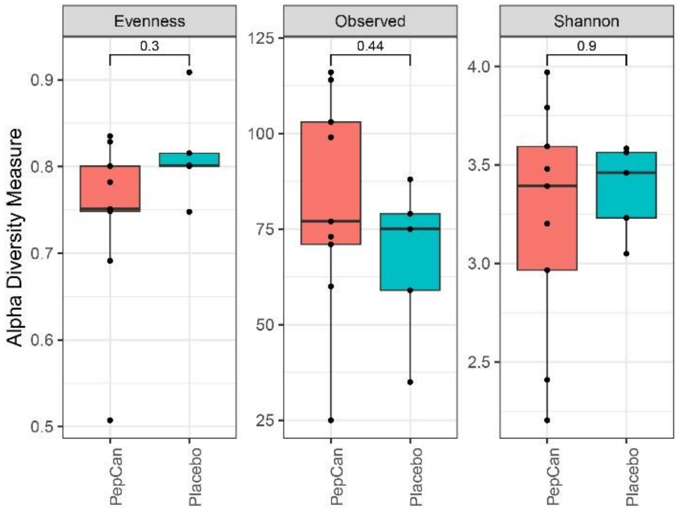

Evenness
Observed
Shannon
0.3
0.44
0.9
Alpha Diversity Measure
0.9
0.8
0.7
0.6
0.5
125
100
75
50
25
4.0
3.5
3.0
2.5
PepCan
Placebo


**D**

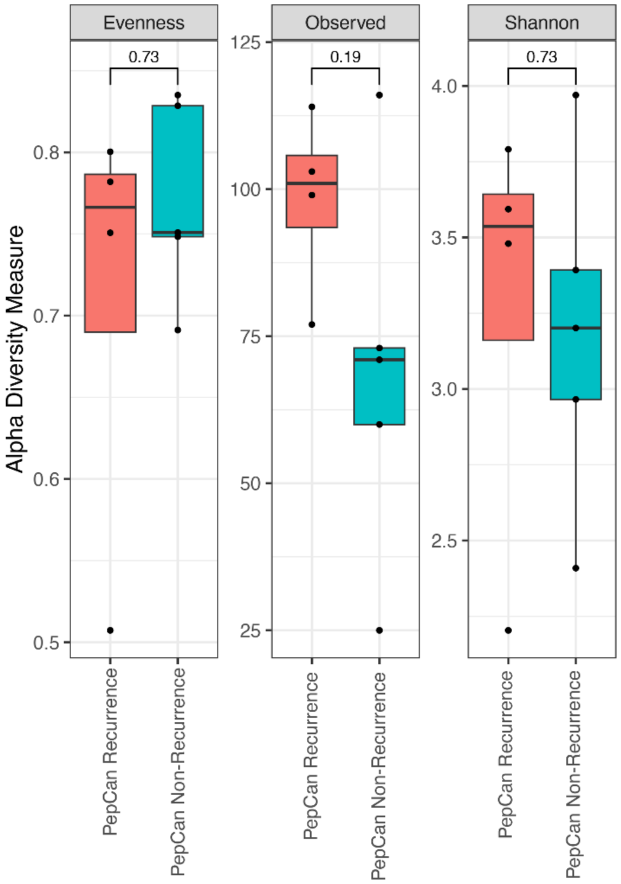

Evenness
Observed
Shannon
0.73
0.19
0.73
Alpha Diversity Measure
0.8
0.7
0.6
0.5
125
100
75
50
25
4.0
3.5
3.0
2.5
PepCan Recurrence
PepCan Non-Recurrence

**Figure S7. Microbiome analyses using amplicon-based rRNA sequencing. A**. Phyla in individual stool samples are shown. Each column represents a stool sample from a particular visit of a particular patient. **B**. Genera in individual stool samples are shown. **C**. Comparisons of alpha diversity metrics between PepCan and placebo groups in pre-vaccination (visit 1) stool samples. No significant differences were found. **D**. Comparisons of alpha diversity metrics between PepCan recurrence and PepCan non-recurrence groups in pre-vaccination (visit 1) stool samples. No significant differences were found. **E**. Phyla in individual oral wash samples are shown. Each column represents a stool sample from a particular visit of a particular patient. **F**. Genera in individual oral wash samples are shown. **G**. Comparisons of alpha diversity metrics between PepCan and placebo groups in pre-vaccination (visit 1) oral wash samples. No significant differences were found. **H**. Comparisons of alpha diversity metrics between PepCan recurrence and PepCan non-recurrence groups in pre-vaccination (visit 1) oral wash samples. No significant differences were found.

**A**

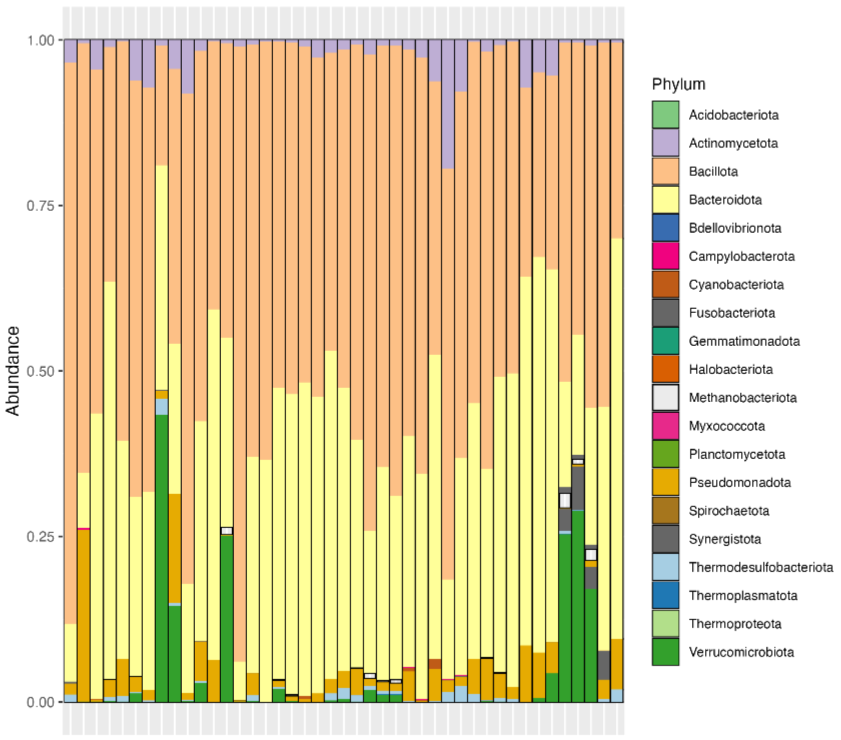

**B**

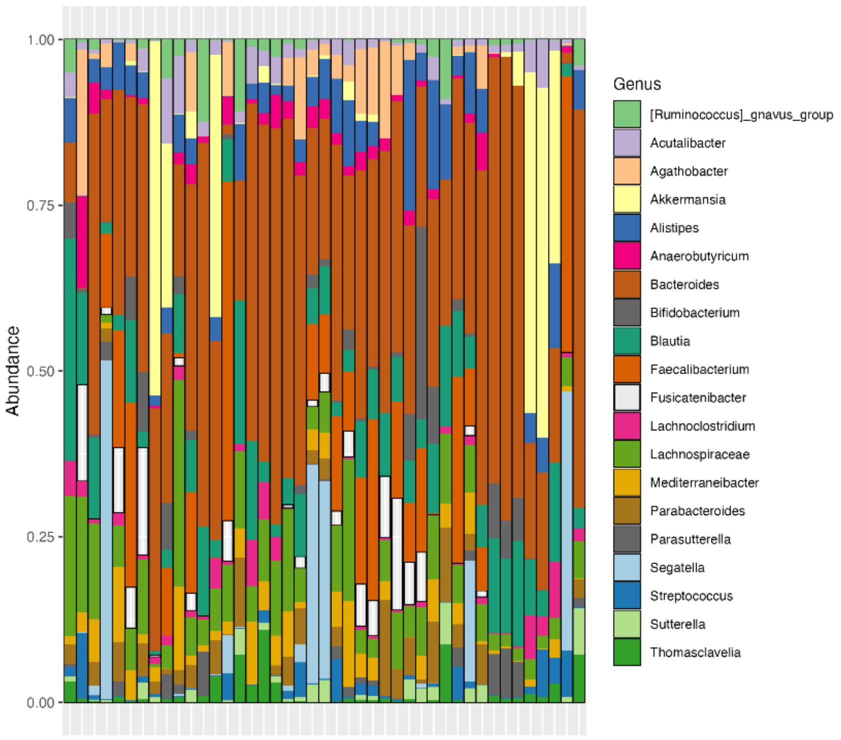
Abundance
1.00
0.75
0.50
0.25
0.00
Genus
[Ruminococcus]_gnavus_group
Acutalibacter
Agathobacter
Akkermansia
Alistipes
Anaerobutyricum
Bacteroides
Bifidobacterium
Blautia
Faecalibacterium
Fusicatenibacter
Lachnoclostridium
Lachnospiraceae
Mediterraneibacter
Parabacteroides
Parasutterella
Segatella
Streptococcus
Sutterella
Thomasclavelia

**C**

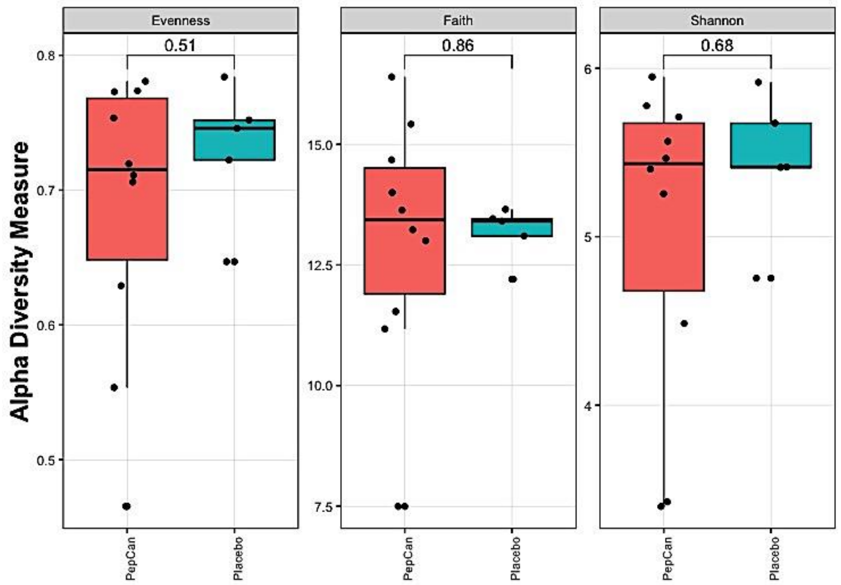
Alpha Diversity Measure
Evenness
0.51
0.8
0.7
0.6
0.5
Faith
0.86
15.0
12.5
10.0
7.5
Shannon
0.68
6
5
4
PepCan
Placebo
PepCan
Placebo
PepCan
Placebo

**D**

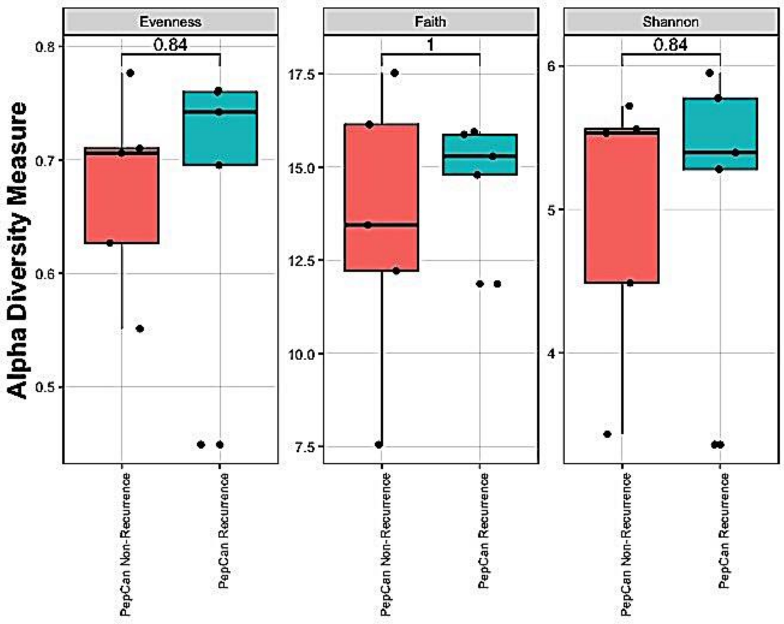
Evenness
Faith
Shannon
0.84
1
0.84
Alpha Diversity Measure
0.8
0.7
0.6
0.5
17.5
15.0
12.5
10.0
7.5
6
5
4
PepCan Non-Recurrence
PepCan Recurrence

**E**

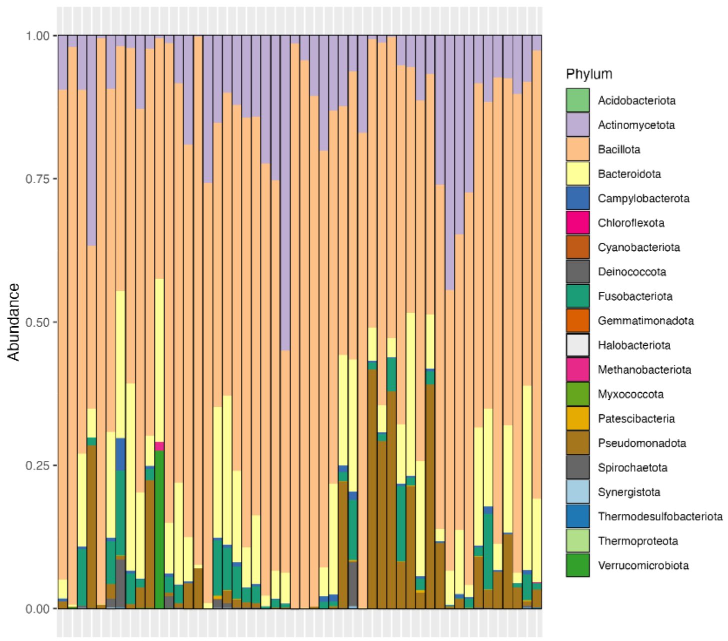
Abundance
1.00
0.75
0.50
0.25
0.00
Phylum
Acidobacteriota
Actinomycetota
Bacillota
Bacteroidota
Campylobacterota
Chloroflexota
Cyanobacteriota
Deinococcota
Fusobacteriota
Gemmatimonadota
Halobacteriota
Methanobacteriota
Myxococcota
Patescibacteria
Pseudomonadota
Spirochaetota
Synergistota
Thermodesulfobacteriota
Thermoproteota
Verrucomicrobiota

**F**

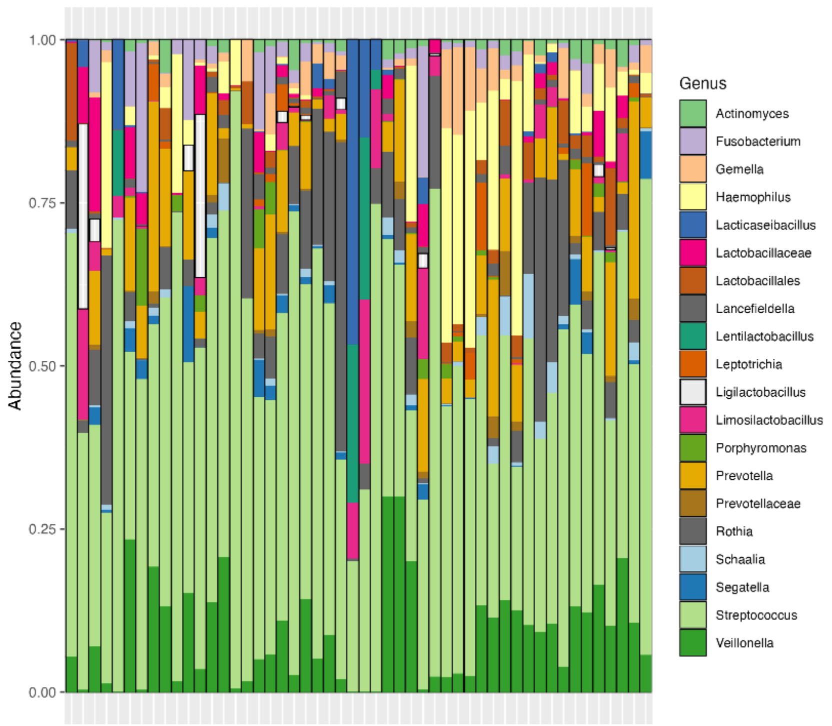
Abundance
1.00
0.75
0.50
0.25
0.00
Genus
Actinomyces
Fusobacterium
Gemella
Haemophilus
Lacticaseibacillus
Lactobacillaceae
Lactobacillales
Lancefieldella
Lentilactobacillus
Leptotrichia
Ligilactobacillus
Limosilactobacillus
Porphyromonas
Prevotella
Prevotellaceae
Rothia
Schaalia
Segatella
Streptococcus
Veillonella

**G**

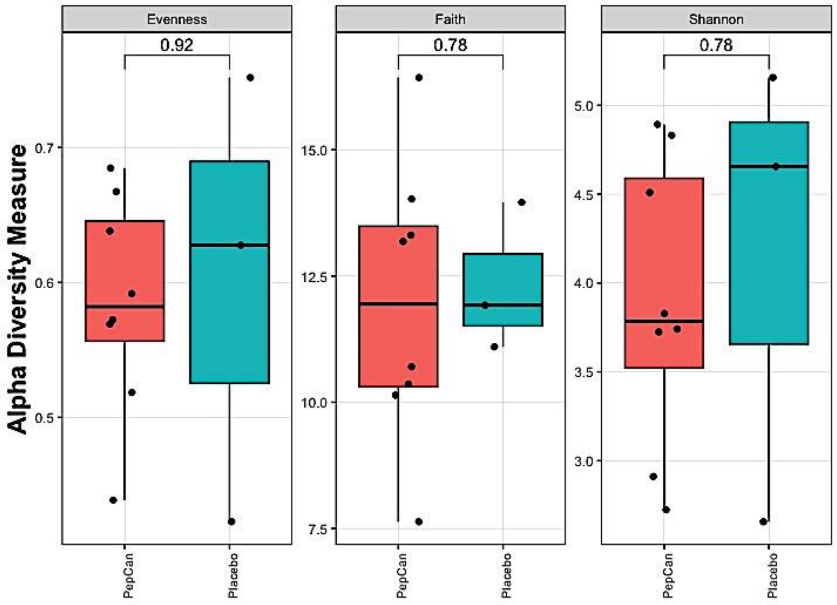
Alpha Diversity Measure
Evenness
0.92
0.7
0.6
0.5
Faith
0.78
15.0
12.5
10.0
7.5
Shannon
0.78
5.0
4.5
4.0
3.5
3.0
PepCan
Placebo
PepCan
Placebo
PepCan
Placebo

**H**

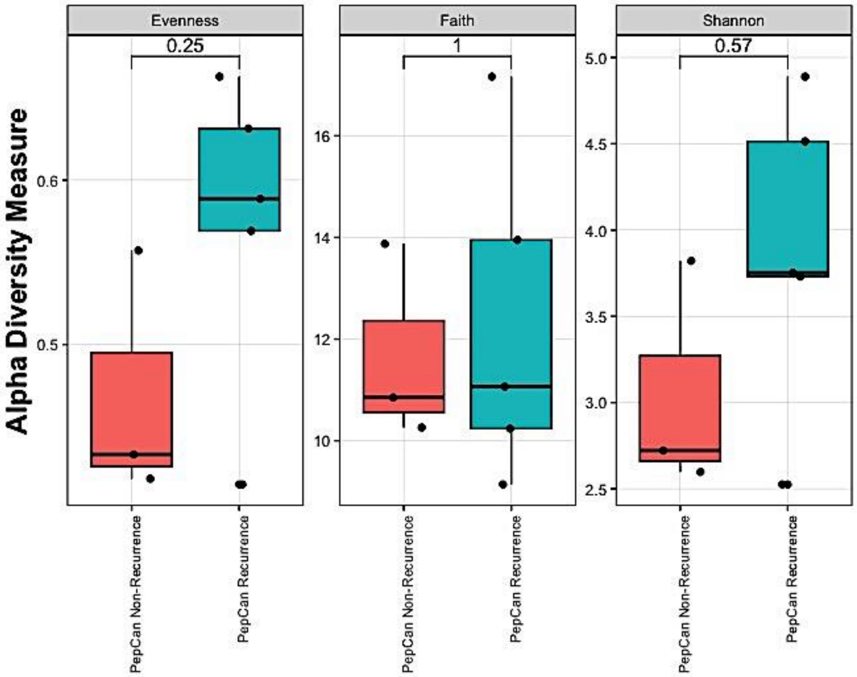
Evenness
Faith
Shannon
0.25
1
0.57
Alpha Diversity Measure
0.6
0.5
16
14
12
10
5.0
4.5
4.0
3.5
3.0
2.5
PepCan Non-Recurrence
PepCan Recurrence
